# ^1^H-MRS metabolites in antipsychotic-responsive versus non-responsive psychosis: a meta- and mega-analysis

**DOI:** 10.64898/2025.12.10.25341974

**Authors:** Bridget King, Kirsten Borup Bojesen, Charlotte Crisp, Andrea de Bartolomeis, Lieuwe de Haan, Camilo de la Fuente-Sandoval, Kara Dempster, Richard J. Drake, Paola Dazzan, Bjørn H. Ebdrup, Lejia Fan, Ariel Graff-Guerrero, Birte Yding Glenthøj, Shiori Honda, Oliver Howes, Li-Chung Huang, Rene Kahn, James MacCabe, Marta Matrone, Kate Merritt, Meghan McIlwain, Philip McGuire, Shinichiro Nakajima, Stephen M. Lawrie, Lena Palaniyappan, Francisco Reyes-Madrigal, Bruce Russell, Akira Sawa, Sukhi Shergill, Krish D. Singh, Iris E. Sommer, James M Stone, Junyu Sun, Sakiko Tsugawa, Fumihiko Ueno, Marieke van der Pluijm, Elsmarieke van de Giessen, James T.R. Walters, Kun Yang, Yen Kuang Yang, Matthew J. Kempton, Alice Egerton

## Abstract

Understanding the mechanisms underlying the response to antipsychotic medications is critical for refining targets for new interventions and predicting clinical outcomes. This study presents a mega-analysis of individual participant data (N = 1,189) from 18 ¹H-MRS datasets to examine differences in neurometabolites in antipsychotic non-responsive compared to antipsychotic-responsive psychosis, accompanied by complementary meta-analyses across the wider published literature (23 studies, N = 1,844). The mega-analysis revealed that antipsychotic non-response is associated with elevated levels of glutamate, Glx (the sum of glutamate and glutamine), choline, and myo-inositol (mI) in the medial frontal cortex (MFC) compared to individuals who showed a good antipsychotic response and healthy controls. Follow-up analyses revealed that elevated MFC Glx in antipsychotic non-responders, compared with responders, is already evident prospectively in first-episode psychosis, whereas elevated mI is most pronounced in individuals meeting criteria for treatment resistance following antipsychotic treatment. The elevations in MFC choline and mI associated with antipsychotic non-response were also detected in the meta-analysis. In both the meta- and mega-analysis, several metabolites were more variable in the patient than the healthy control groups. Collectively, these data provide the most robust evidence to date linking antipsychotic non-response in psychosis to elevations in medial frontal glutamate, choline and mI. The findings support the continued investigation of glutamate-acting and inflammatory pathway-associated interventions for psychosis and schizophrenia, and particularly for patients who have not responded to antipsychotic treatment.

## Introduction

The neurobiological mechanisms underlying the clinical response to antipsychotic medication in psychosis is unclear. Up to 30% of patients do not respond adequately to conventional antipsychotics and meet criteria for treatment resistant schizophrenia (TRS) (1,2). Understanding the neurobiological mechanisms that contribute to a poor antipsychotic response and identifying biomarkers that can predict response could potentially help guide clinical interventions and/or inform new therapeutics. First-line antipsychotics primarily target dopamine D2 receptors and positron emission tomography studies indicate striatal dopaminergic tone plays a role in antipsychotic response (3). However, novel therapeutic development for antipsychotic non-responsive schizophrenia likely requires identification of non-dopaminergic targets.

Proton magnetic resonance spectroscopy (^1^H-MRS) can measure several different brain metabolites relevant to schizophrenia pathophysiology. In addition to glutamate (reported alone or in combination with glutamine as Glx) and GABA, ¹H-MRS can quantify several other metabolites, including N-acetylaspartate (NAA; a marker of neuronal integrity), myo-inositol and choline-containing compounds (mI, Cho; markers of astroglial activation and neuroinflammation, with Cho also involved in membrane metabolism), and glutathione (GSH; an intracellular antioxidant) (4).

Recent meta-analyses of group mean differences have indicated that ¹H-MRS metabolites in the medial frontal cortex (MFC) may vary according to antipsychotic treatment response. Nakahara et al. (5) found that, compared with healthy controls, TRS was associated with elevated glutamate or Glx, in the midcingulate cortex (MCC), a subregion of the MFC, while glutamate was decreased in the MCC in non-TRS patients. This is consistent with our previous mega-analysis finding of higher MFC glutamate levels among patients with more severe symptoms (6). Additionally, while a meta-analysis by Smucny et al. (7) reported no significant differences in MFC or striatal glutamate levels between TRS, non-TRS, and healthy controls, MFC choline and mI were increased in TRS compared to both non-TRS and healthy controls. Together, these findings suggest a potential role for more severe glutamatergic dysregulation, astroglial activation and neuroinflammatory processes in antipsychotic non-responsive illness.

Previous analyses of ^1^H-MRS metabolites in relation to antipsychotic response (5,7) relied on group-level data. In contrast, by collating individual participant data from different studies, mega-analytic approaches offer increased statistical power, more accurate effect size estimation, and a better ability to test for potential moderating variables. Furthermore, while previous meta-analyses have focused on cross-sectional data, a key question is whether differences in metabolites according to treatment response outcomes are also detectable prospectively, from first-episode psychosis, so that they could be used to predict antipsychotic response. This has been suggested by some (8–13), but not all individual studies (14–16). Glutamatergic metabolites in the MFC, dorsolateral prefrontal cortex (DLPFC) and thalamus (17), as well as GSH in the MFC (18), all show more variability in schizophrenia than in controls, suggesting biological heterogeneity between patients. In the MFC and basal ganglia, glutamate variability was greatest among the most symptomatic patients (17), which could suggest more severe disturbance of glutamate regulatory mechanisms. However, it remains unknown whether variability differs according to antipsychotic response. This question is also important, as biomarkers with lower variability provide more stable and consistent biomarkers of treatment response.

In the current study we aimed to comprehensively evaluate the profile of ^1^H-MRS metabolites in relation to treatment response in schizophrenia by conducting a mega-analysis of individual participant-level data. We hypothesised that a) in the MFC, glutamate, Glx, Cho and mI would be elevated in antipsychotic non-responders compared in antipsychotic responders and healthy controls; b) in the MFC and basal ganglia, variability in glutamate metabolites would be greater in antipsychotic non-responders than responders and controls. We further examined whether differences in metabolite levels according to treatment response are detectable prospectively, by focusing on studies in which ¹H-MRS measures were acquired in first-episode psychosis with minimal prior antipsychotic exposure, to assess whether baseline metabolites are associated with subsequent response. In addition, we investigated whether group differences in metabolites were specific to TRS as opposed to broader definitions of antipsychotic non-response. We additionally conducted a meta-analysis of the wider literature to examine group differences in standardised mean differences and variability.

## Methods

### Search strategy and study selection

The study was conducted according to PRISMA guidelines and registered on PROSPERO prior to search (CRD42022346139). Web of Science database searches identified journal articles published between January 1, 1980, and August 10, 2024, using the following search terms: (schizophreni* OR psychosis) AND (glutamate OR glutamine OR glutamatergic OR Glx OR N-acetylaspartate OR NAA OR myo-inositol OR glutathione OR GSH OR creatine OR choline) AND (MRS OR magnetic resonance spectroscopy). For the meta-analysis, an updated search was conducted in November 2025 (Supplementary Figure 1). Study inclusion required reporting of *in vivo* brain ^1^H-MRS metabolite values in individuals with schizophrenia, first-episode psychosis or related disorders, classified according to their response to antipsychotic treatment. We kept the original definitions of antipsychotic treatment response and non-response used in each study (Supplementary Table 2). This differed between studies but commonly involved achieving a minimum percentage reduction on symptom scales or meeting established remission criteria. Non-response was defined conversely to response and, in some studies, aligned with consensus criteria for TRS. Studies not categorising patients according to antipsychotic treatment response were excluded. Screening and selection of published studies was performed independently by two authors (B.K, J.S) in Rayyan (19). Searches for relevant grey literature were conducted in Medrxiv and OpenGrey. Any discrepancies were resolved through discussion.

### Individual patient data requests – mega-analysis

Authors of the 21 studies identified before August 2024 that met our inclusion criteria were contacted at least twice to request anonymised participant-level data on ¹H-MRS metabolite levels (Glu, Glx, NAA, Cho, mI, GABA and GSH). Data transfer was subject to institutional data sharing agreements, which typically required 3-6 months to establish. For longitudinal studies, only the ^1^H-MRS data for the first time point were included, and response was determined at the final follow-up timepoint. We also requested participant-level clinical and demographic information, including age, sex, duration of illness, antipsychotic medication dose and Positive and Negative Syndrome Scale (PANSS) symptom severity scores.

### Data extraction – meta-analysis

Metabolite means and standard deviations of treatment responders and treatment non-responders with psychosis and healthy controls from the published literature were extracted and categorised into the MFC, DLPFC, thalamus and basal ganglia. Further details are presented in the supplementary materials.

### Statistical analyses

Analyses were conducted in R (version 4.3.0). All analyses were restricted to variables for which a minimum of 3 independent datasets were available. We conducted separate analyses for each ^1^H-MRS metabolite due to their biologically distinct functions, and brain region due to expected regional differences. P < 0.05 was considered statistically significant for comparisons for which we had *a priori* hypotheses (group differences in MFC glutamate, Glx, Cho and mI; and variability of glutamate metabolites in MFC and basal ganglia). Benjamini-Hochberg false discovery rate (FDR) correction applied to all other comparisons, using a Q threshold of 10%.

#### Mega-analysis of individual patient data

Group differences in individual patient data were determined using linear mixed models using the lmer and emmeans packages (20,21). As variability between groups may differ, heteroscedastic and homoscedastic models were compared and the Akaike information criterion (AIC) and likelihood ratio tests (LRT) were used to select the best-fitting model. Once the variance structure was established, group differences were assessed using linear mixed models, with metabolite as the dependent variable, group as a fixed effect (responder, non-responder, healthy controls), and study as a random effect, covarying for age and sex. Significant group effects were followed up with post hoc pairwise comparisons, with effect sizes expressed as Glass’s Δ. Glass’s Δ was calculated using the standard deviation of the reference group for each comparison: for non-responder versus responder comparisons, the responder group standard deviation was used, and for comparisons involving healthy controls, the healthy control group standard deviation was used.

We conducted two sub-group analyses as part of the secondary analyses. First, we restricted the sample to prospective studies in first-episode psychosis. Second, we further subdivided the non-responder group to examine treatment resistance more specifically, by introducing separate groups for non-responders who did or did not meet TRS criteria as separate levels in the model, resulting in four groups (TRS, non-TRS, responders, and healthy controls).

We also conducted a sensitivity analysis limited to studies which used CSF correction to quantify metabolite levels. Follow-up analyses examined whether group differences in metabolites were influenced by medication dose (CPZ) or symptom severity (PANSS total scores) by including either a group x CPZ dose or a group x PANSS score interaction term in the model.

Where the heteroscedastic models provided a better fit, we examined whether the residual variance differed significantly between groups. We used the glmmTMB R package (22) to fit models that estimated mean structures (as in our primary linear mixed models) and with an additional dispersion model that allowed the residual variance to vary by group. Pairwise contrasts of these dispersion parameters (the estimated log-residual variances for each group) were then performed using Wald tests, which tests whether the difference in log-variances is significantly different from zero.

#### Meta-analyses of mean differences and variability

Meta-analyses of standardised mean differences (SMDs) between groups were conducted using random-effects models using the “metafor” R package (23). SMDs were calculated using the Hedges’ g statistic and heterogeneity using the I^2^ value. Further details are provided in the supplementary materials. Meta-analyses of variability were quantified using the log coefficient variation ratio (CVR), as previously described (17,24).

## Results

### Mega-analysis of individual patient data

A total of 18 out of 21 eligible studies contributed individual participant data (Supplementary Figure 1 and Supplementary Table 1). This resulted in a dataset of 1,189 participants, including 476 patients defined as treatment non-responders, 427 patients defined as treatment responders, and 286 healthy controls. Groups did not differ in sex distribution, but age was higher in the non-responder compared to responder and control groups. Antipsychotic CPZ dose and PANSS total scores were higher in the non-responder compared to the responder group (P <.001) (Supplementary Table 3). Voxel placements in the MFC varied by study, an illustration is presented in Supplementary Figure 2.

### Primary linear mixed models

Except for GABA in the MFC, LRT and AIC revealed significantly better model fit for heteroscedastic models, indicating that the groups had unequal variance in all other metabolites in all other regions (Supplementary Table 4).

There were significant overall effects of group for Glu, Glx, Cho, and mI in the MFC (Table 1). Subsequent post hoc analysis found the non-responder group had significant elevations in MFC Glu (Estimate (E) = 0.340, df = 852, P = 0.016, Glass’s Δ = 0.21), Glx (E =0.606, df =690, P =0.002, Glass’s Δ = 0.29), Cho (E = 0.075, df = 634, P = 0.028, Glass’s Δ = 0.22), and mI (E =0.355, df = 586, P =0.001, Glass’s Δ = 0.35) compared to the responder group. The elevations in MFC Glu (E = 0.360, df = 852, P = 0.026, Glass’s Δ = 0.22), Glx (E = 0.627, df = 690, P = 0.005, Glass’s Δ = 0.30), Cho (E = 0.171, df = 634, P<.001, Glass’s Δ = 0.51), and mI (E = 0.623, df = 586, P<.001, Glass’s Δ = 0.61) in the non-responder group were also significant compared to controls. In addition, MFC Cho (E = 0.096, df = 634, P = 0.005, Glass’s Δ = 0.51) and mI (E = 0.269, df = 586, P = 0.026, Glass’s Δ = 0.26) were significantly elevated in the responder group compared to controls (Figure 1, Table 2).

**Figure 1.**
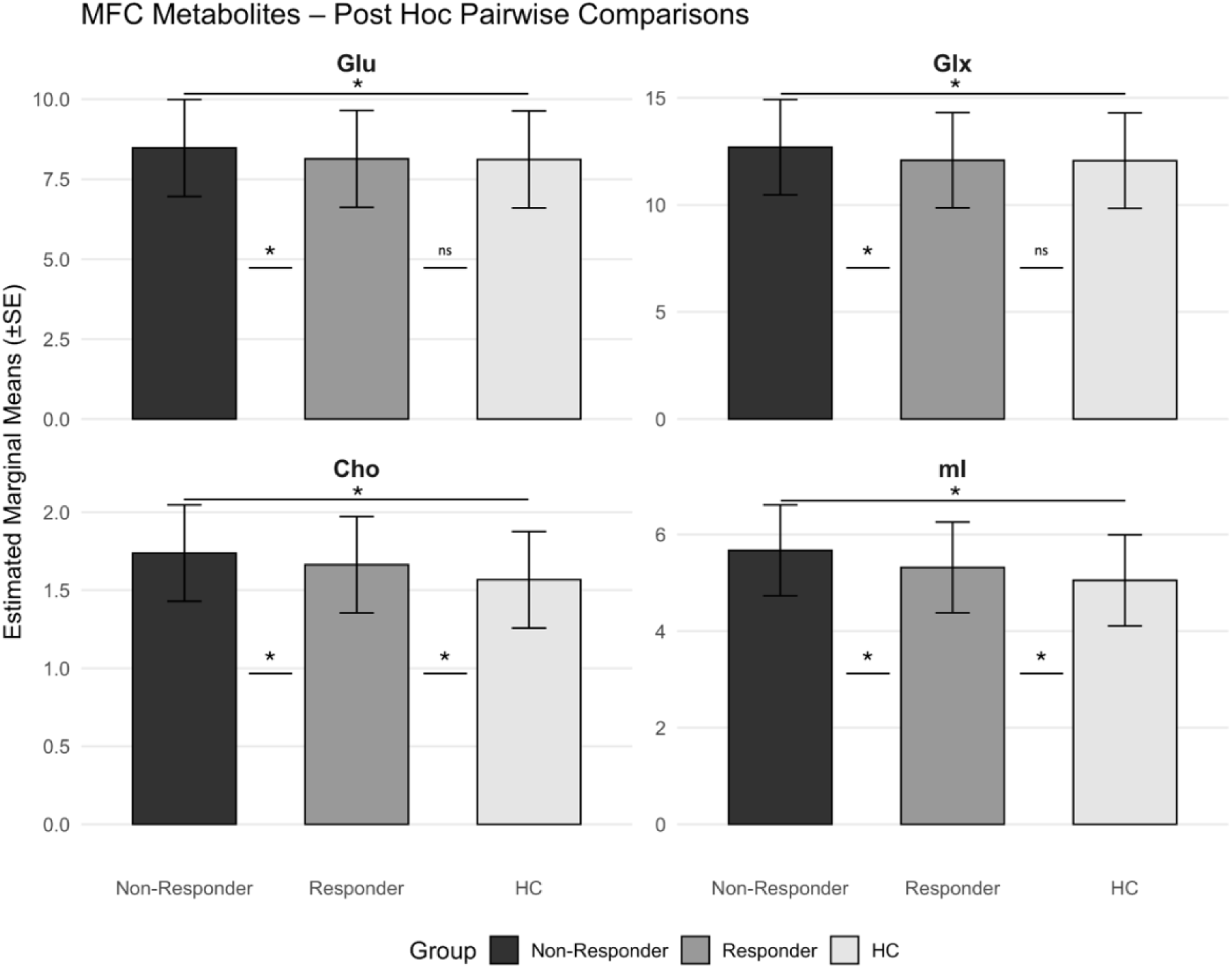
Bar graphs displaying estimated marginal means with standard error (±SE) for MFC Glu, Glx, Cho and mI for the non-responder, responder and healthy control (HC) groups. Significant pairwise comparisons are denoted by an asterisk (*), non-significant pair-wise comparisons are indicated by ‘ns’. Abbreviations: Cho: choline; Glu: glutamate; Glx: glutamate+glutamine; MFC: medial frontal cortex; mI: myo-inositol.

**Table 1:** Individual patient data – ^1^H-MRS metabolite group effects.

| Metabolite | Fixed effect | N cohorts | N participants | df | F | P |
| --- | --- | --- | --- | --- | --- | --- |
| **Glu** |  |  |  |  |  |  |
| MFC | Group (NR, R, HC) | 20 | 876 | 852 | 3.387 | **0.034** |
| DLPFC | Group (NR, R, HC) | 3 | 187 | 180 | 5.210 | 0.006 |
| Thalamus | Group (NR, R, HC) | 8 | 253 | 243 | 1.340 | 0.264 |
| Basal Ganglia | Group (NR, R, HC) | 9 | 310 | 297 | 0.748 | 0.474 |
| **Glx** |  |  |  |  |  |  |
| MFC | Group (NR, R, HC) | 17 | 711 | 690 | 5.815 | **0.003** |
| Thalamus | Group (NR, R, HC) | 7 | 216 | 207 | 0.298 | 0.743 |
| Basal Ganglia | Group (NR, R, HC) | 9 | 311 | 298 | 0.750 | 0.473 |
| **NAA** |  |  |  |  |  |  |
| MFC | Group (NR, R, HC) | 17 | 696 | 675 | 5.622 | 0.003 |
| DLPFC | Group (NR, R, HC) | 3 | 187 | 180 | 4.468 | 0.013 |
| Thalamus | Group (NR, R, HC) | 8 | 261 | 251 | 0.998 | 0.370 |
| Basal Ganglia | Group (NR, R, HC) | 9 | 312 | 299 | 0.858 | 0.425 |
| **Cho** |  |  |  |  |  |  |
| MFC | Group (NR, R, HC) | 16 | 654 | 634 | 11.518 | **< .001** |
| Thalamus | Group (NR, R, HC) | 7 | 221 | 212 | 1.062 | 0.348 |
| Basal Ganglia | Group (NR, R, HC) | 8 | 268 | 256 | 0.552 | 0.577 |
| **mI** |  |  |  |  |  |  |
| MFC | Group (NR, R, HC) | 15 | 605 | 586 | 12.783 | **< .001** |
| Thalamus | Group (NR, R, HC) | 7 | 218 | 208 | 1.552 | 0.214 |
| Basal Ganglia | Group (NR, R, HC) | 8 | 270 | 258 | 0.328 | 0.721 |
| **GABA** |  |  |  |  |  |  |
| MFC | Group (NR, R, HC) | 4 | 253 | 246 | 0.274 | 0.760 |
| **GSH** |  |  |  |  |  |  |
| MFC | Group (NR, R, HC) | 3 | 158 | 151 | 1.110 | 0.332 |

Significant results (after FDR correction as described in the methods) are indicated in bold. For studies that acquired data at multiple sites, each site was treated as a separate cohort. Abbreviations: Cho: choline; DLPFC: dorsolateral prefrontal cortex; GABA: gamma-aminobutyric acid; Glu: glutamate; Glx: glutamate+glutamine; GSH: glutathione; HC: healthy controls; mI: myo-inositol; MFC: medial frontal cortex; NAA: N-acetylaspartate; NR: non-responder group; R: responder group.

**Table 2:**
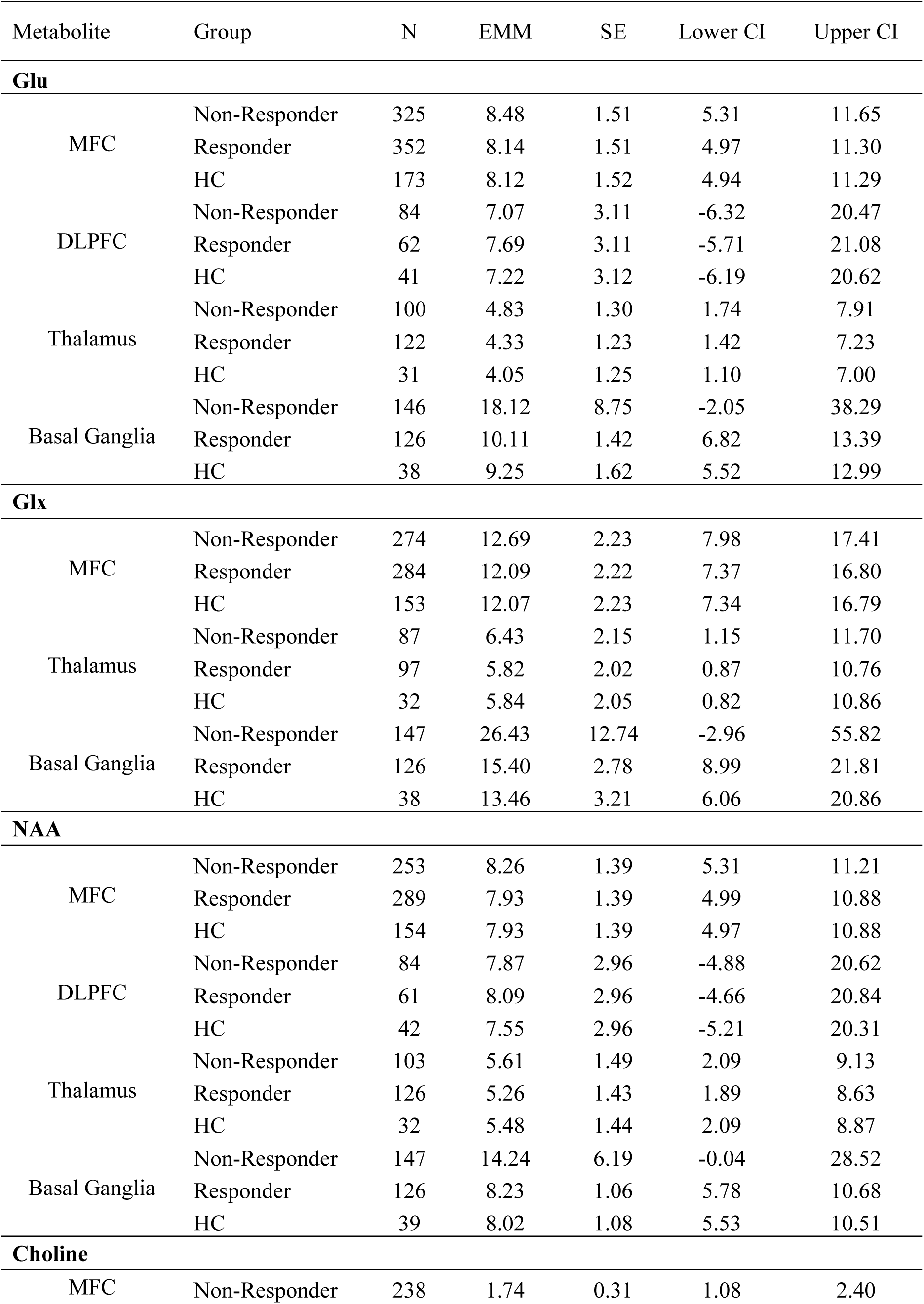
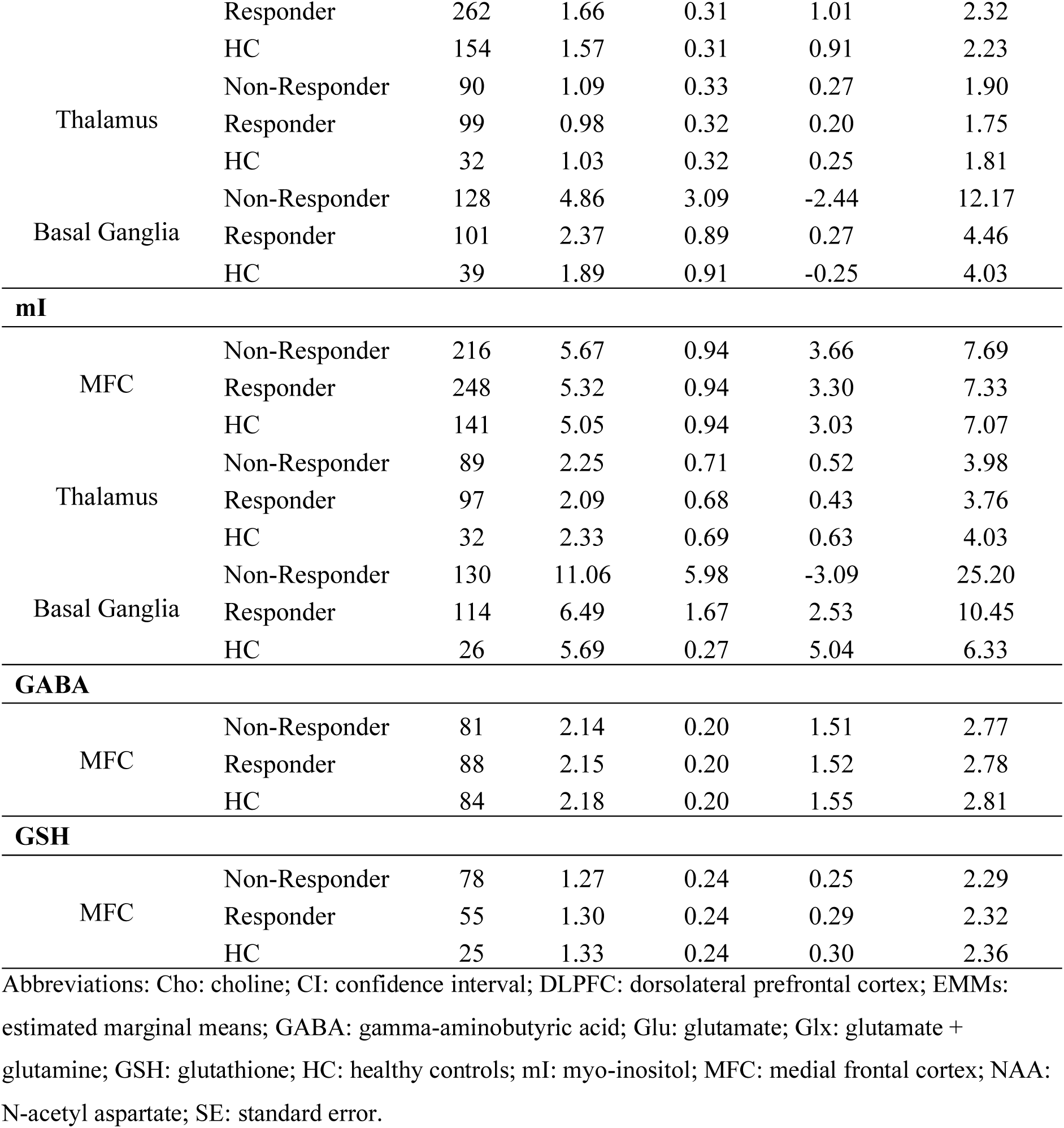
Estimated marginal means (EMMs) for ^1^H-MRS metabolites in the treatment non-responder, responder and healthy control groups.

Metabolite distributions in the DLPFC and basal ganglia showed evidence of non-normality. To address this, we applied log transformations and re-ran the linear mixed models as a sensitivity analysis; the results of these models are presented in Supplementary Table 5.

### Subgroup analyses

#### Prospective studies in first-episode psychosis

When the analysis was limited to prospective studies in first-episode psychosis, the linear mixed models indicated significant group effects for Glx (F = 4.60, df = 335, P = 0.010), NAA (F = 3.188, df = 350, P = 0.042) and Cho (F = 4.265, df = 310, P = 0.014) in the MFC. Post hoc comparisons revealed elevations in MFC Glx (Estimate (E) = 0.824, df = 355, P = 0.002, Glass’s Δ = 0.41) and NAA (E = 0.379, df = 350, P = 0.009, Glass’s Δ = 0.34) in patients who would later be defined as non-responders compared to responders. MFC Cho was elevated in patients later defined as either responders (E = 0.100, df = 310, P = 0.017, Glass’s Δ = 0.36) or non-responders (E = 0.159, df = 310, P = 0.003, Glass’s Δ = 0.57) compared to controls. Further statistics are presented in Supplementary Table 6.

#### Treatment resistant samples

Models examining the TRS status of individuals within the non-responder group (TRS vs. non-TRS) returned significant effects of TRS status on MFC NAA (F = 4.03, df = 674, P = 0.007), Cho (F = 8.61, df = 633, P < .001), and mI (F = 12.02, df = 585, P < .001). Post hoc comparisons found MFC mI was significantly elevated in those defined as TRS relative to all other groups: those non-responsive but below the TRS threshold (E = 0.654, df = 585, P = 0.001, Glass’s Δ = 0.64), responders (E = 0.788, df = 585, P < .001, Glass’s Δ = 0.78), and controls (E = 0.985, df = 585, P < .001, Glass’s Δ = 0.97). The TRS group also showed elevations in MFC Cho (E = 0.090, df = 687, P = 0.039, Glass’s Δ = 0.33) and NAA (E = 0.302, df = 674, P = 0.029, Glass’s Δ = 0.23) compared to the responder group, and NAA (E = 0.332, df = 674, P = 0.010, Glass’s Δ = 0.26) and Cho (E = 0.200, df = 633, P < .001, Glass’s Δ = 0.59) compared to the control group. Further statistics are presented in Supplementary Table 7.

#### Relationships with antipsychotic dose and symptom severity

CPZ dose was not significantly associated with regional metabolite levels across the whole patient sample. There was a significant CPZ × group interaction for MFC mI (t (366) = 2.17, p = 0.031), whereby CPZ dose was positively associated with mI in the non-responder group but not in the responder group. In the non-responder group, each 100-unit increase in CPZ dose was associated with a +0.071 change in mI (95% CI: 0.011–0.130), whereas the slope was close to zero in the responder group (−0.031, 95% CI: −0.110 to 0.048). PANSS total scores were not significantly associated with any regional metabolite levels and there were no significant interactions between treatment response status and PANSS total scores on metabolite levels.

#### Group differences in metabolite variation

The non-responder group showed significantly greater variation in MFC Cho, and in thalamic Glu, Glx, Cho, and mI compared to the responder group. Both the non-responder and responder groups showed significantly greater variation relative to controls in MFC Glu, NAA, Cho, and GSH, as well as in thalamic NAA and Cho. The non-responder group also showed increased variation in thalamic Glu and mI, and the responder group showed increased variation in basal ganglia Cho compared with controls (Table 3).

**Table 3:**
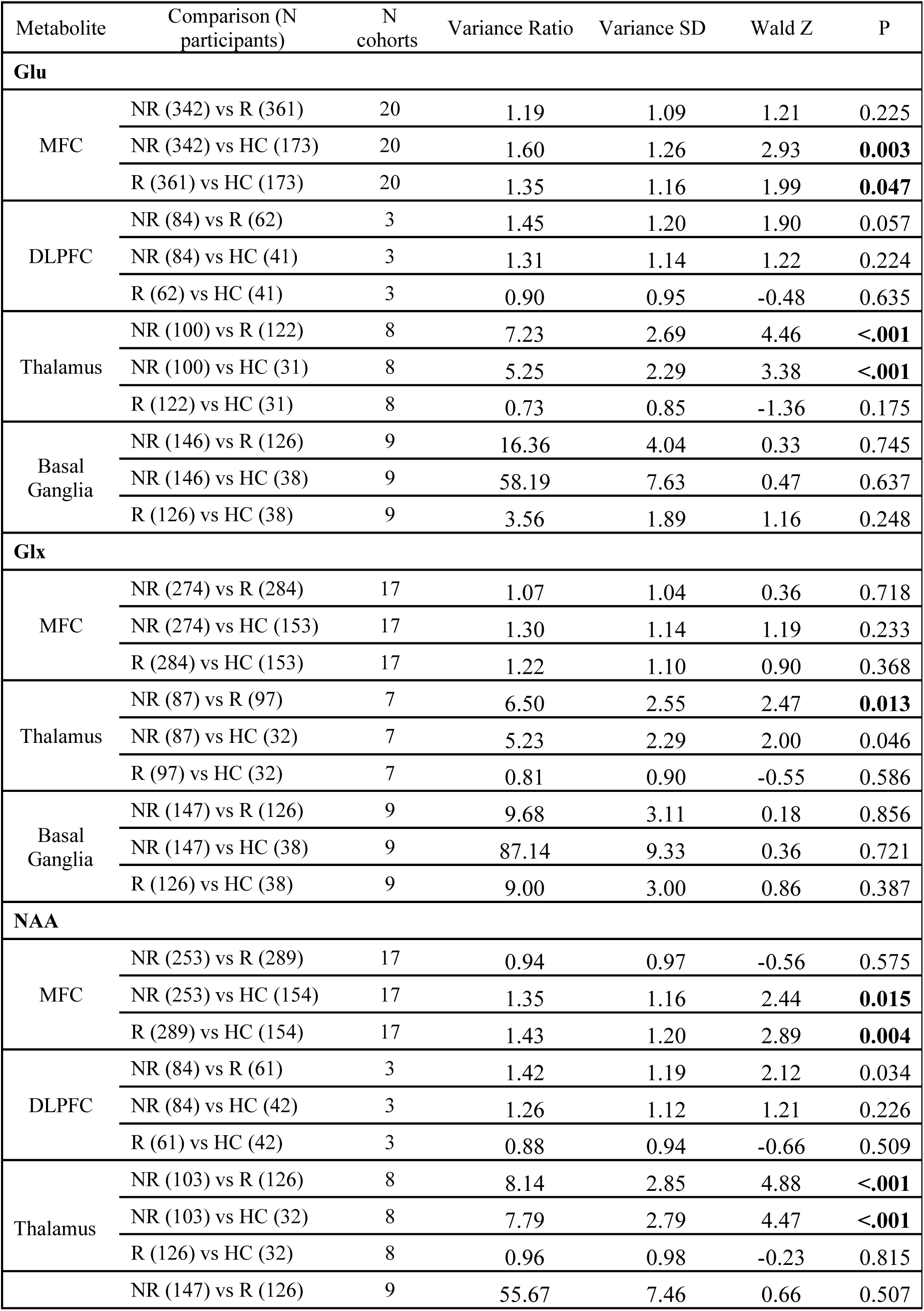
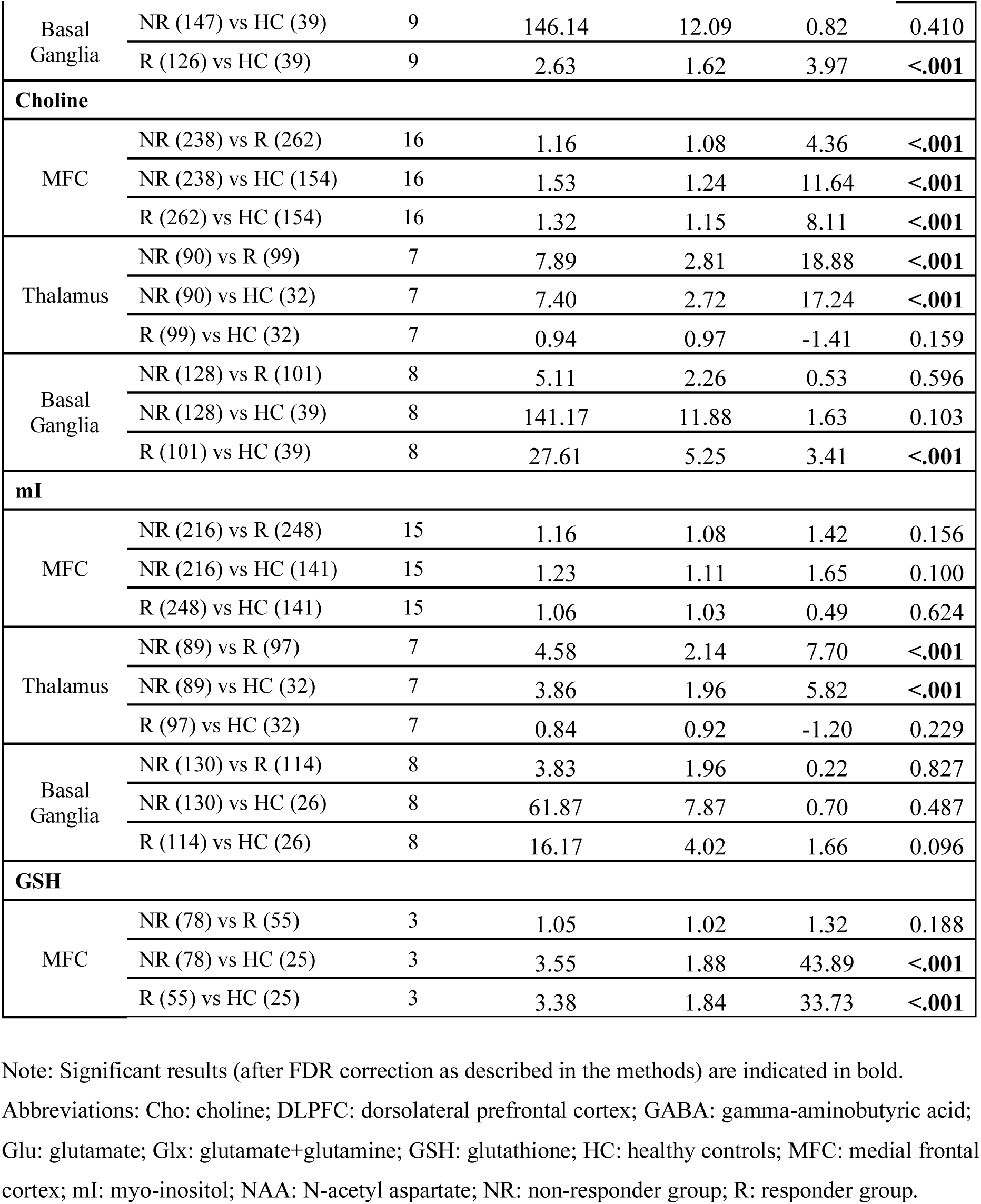
Pairwise comparisons of metabolite variation in the treatment non-responder, responder and healthy control groups.

#### Meta-analysis of mean differences

The meta-analysis of the published literature included 23 studies comprising 1,844 participants, representing an additional five studies and 655 participants beyond those included in the mega-analysis (Supplementary Figure 1 and Supplementary Table 9).

Consistent with the mega-analysis, the meta-analysis also showed elevated MFC Cho and mI in the non-responder group compared with both the responder and control groups, and higher MFC Cho in the responders compared with controls. The elevations in MFC Glu and Glx in the non-responder compared to responder and control groups detected in the mega-analysis were not significant in the meta-analysis, and no further significant overall group differences in metabolite levels were apparent (Supplementary Figure 3 and Supplementary Table 10).

#### Meta-analysis of variability (CVR)

Consistent with the mega-analysis, the meta-analysis of variability also demonstrated that the non-responder group showed greater variability in MFC Glu, Glx, and NAA compared to controls. The significant group differences in MFC Cho variability detected in the mega-analysis were not significant in the meta-analysis (Supplementary Figure 4). In the meta- (but not mega-) analysis, the non-responder group showed significantly greater variability in DLPFC Glu compared to the responder group, and greater variability in thalamic Glx compared to controls. In the basal ganglia, the responder group showed lower variability in Glu and higher variability in NAA relative to controls (Supplementary Table 11).

## Discussion

The current manuscript presents a mega-analysis of ^1^H-MRS metabolites in 1,189 individuals from 18 studies in relation to antipsychotic response in schizophrenia, supplemented with meta-analyses across the entire published literature (23 studies and 1,844 individuals). In line with our hypotheses, analysis of the individual participant data revealed elevations in MFC Glu and Glx (glutamate + glutamine), choline, and mI in the antipsychotic non-responder group compared to both the responder and healthy control groups. Moreover, the elevations in MFC Glx in the non-responder group were present in studies employing prospective designs, indicating they are present early in illness and may predict subsequent non-response. Elevations in MFC glutamatergic metabolites associated with non-response did not differ according to whether individuals in the non-responder group were classified as TRS. In contrast, mI levels in the MFC were significantly higher in TRS samples, compared to all groups. By leveraging participant-level data across studies, this analysis offers greater statistical power and precision compared with meta-analytic approaches. It thereby provides more robust evidence that elevations in medial frontal glutamate metabolites and markers of astroglial activation and neuroinflammation are associated with antipsychotic non-response in psychosis.

The elevations in MFC Glu and Glx levels in the antipsychotic non-responder compared with both the responder and control groups in the individual patient data were associated with small to moderate effect sizes (Glass’s Δ Glutamate: 0.21 Glx: 0.29), and did not reach significance in the current or previous meta-analyses (7). This demonstrates the increased statistical power of mega-analytic techniques to detect smaller effect sizes. The elevations in MFC Glx, but not glutamate, in the non-responder group were detected prospectively. As elevations in Glx across the whole sample showed slightly larger effect sizes, and variability in MFC glutamate, but not Glx, was greater within both patient groups compared to controls, MFC Glx may provide a somewhat more consistent marker of non-response than MFC glutamate. This difference may also relate to the limited ability to separate glutamate and glutamine signals at lower MRI field strengths. The elevations in glutamate or Glx linked to antipsychotic non-response did not extend to other brain regions. In the DLPFC, there was some indication that the responder rather than the non-responder group showed glutamate elevations; however, this result was based on a small number of studies and did not survive correction for multiple comparisons. No group differences in glutamate or Glx levels were detected in the thalamus and basal ganglia. Our results therefore indicate a regional specificity of MFC glutamate and Glx elevation in relation to antipsychotic non-response, with elevations in MFC Glx also being detectable prospectively. These findings could suggest that compounds that reduce glutamate levels or stabilise glutamate signalling may be beneficial for patients who show a limited or no response to conventional antipsychotic treatment.

MFC choline and mI were highest in the TRS group, followed by the non-responder, responder, then control groups. This indicates that these markers of membrane turnover or breakdown (choline) (25), and astroglial activation or neuroinflammation (mI) (4,26,27) are overall elevated in schizophrenia (28,29), and that the extent of this elevation scales with a poor response to antipsychotic treatment. Elevations in MFC choline and mI within the non-responder compared to the responder and control groups were also detected in our meta-analysis. They are consistent with findings from a previous meta-analysis (7) showing increased choline and mI in TRS, and our results indicate that these elevations extend to a broader group of poor responders earlier in illness or not meeting TRS criteria. The elevations in MFC mI in the non-responder compared to the responder group showed a slightly larger effect size (Glass’s Δ = 0.35) than was detected for MFC glutamate or Glx (0.21 and 0.29, respectively), whereas the effect size for MFC choline was similar (0.22).

MFC choline levels also showed significantly greater variability in patient groups, whereas mI levels did not, suggesting mI may be a more consistent and pronounced marker of antipsychotic non-response. However, mI levels in the non-responder group were positively associated with antipsychotic dose, and mI elevations were not observed in prospective designs. This suggests that increased mI in the non-responder group may partly reflect the effects of antipsychotic exposure, including the higher doses prescribed to patients showing a poor response (see reference (30) for further discussion). It is important to acknowledge the ambiguity in this relationship: while our prospective findings support a medication effect, it remains plausible that elevated mI could be a primary marker of a more severe pathophysiology that both fails to respond to treatment and elicits higher prescribed medication doses. In contrast, elevations in choline were present in both non-responders and responders compared to controls within prospective studies. The ^1^H-MRS choline signal is thought to reflect membrane turnover, as well as being a marker of neuroinflammation and astroglial activation (similar to mI), as both metabolites are found in higher concentrations in glia than in neurons (31). Overall, these results suggest that individuals with psychosis who have not shown a good response to antipsychotic treatment may benefit from therapeutics targeting neuroinflammatory mechanisms. Anti-inflammatory agents have shown some efficacy in reducing symptoms when used as an adjunct to antipsychotic treatment (32); however, their effectiveness in individuals who do not respond to antipsychotics specifically remains to be established.

Neither the mega-nor meta-analysis detected group differences in GABA or GSH levels in relation to antipsychotic response. However, there were a lower number of studies available for this analysis, and for GSH there was also evidence of greater variability within the patient compared to control groups, as previously reported in a wider case-control meta-analysis (18). Both factors may have reduced power to detect mean group differences. There was some indication of an overall group difference in MFC NAA levels in relation to response, but this was not hypothesis driven and did not survive multiple comparisons correction.

### Strengths and limitations

This collaborative mega-analysis, comprising 1,189 individuals from 18 datasets, enabled a more powerful and nuanced investigation of glutamate, choline, and mI in relation to antipsychotic response than prior meta-analyses. It incorporates data drawn from 12 countries and encompassing different stages of illness. We supplemented the mega-analysis with meta-analyses of standardised mean differences and variability, which alone represent the largest meta-analyses of ^1^H-MRS studies and antipsychotic response in psychosis and schizophrenia to date.

The study also has some limitations. Patient cohorts, ^1^H-MRS data acquisition and analysis protocols differ between individual studies, and our analysis only partially addressed these differences. Antipsychotic response was measured with several instruments and variably defined across individual studies, including thresholds on standardised rating scales (PANSS, BPRS, CGI); meeting / not meeting remission criteria; or meeting / not meeting criteria for TRS, which adds heterogeneity. Our analysis did not account for further patient subgroups, including ultra treatment resistant schizophrenia. Antipsychotic response may fluctuate but was only defined at one timepoint, and we did not investigate longitudinal within-subject changes in ^1^H-MRS metabolites in relation to treatment response. Although our individual patient data represents 18 datasets from the 21 eligible studies, the absence of data from the remaining studies may have introduced selection bias and reduced generalisability. However, the main findings of elevated MFC choline and mI in non-responders were also present in our meta-analysis of the entire literature. While our results present robust, well-powered evidence linking glutamatergic (glutamate and Glx) and neuroinflammatory (choline and mI) mechanisms to a poor antipsychotic response, the effect sizes for group differences as observed with ^1^H-MRS are in the small to moderate range. Therefore, while these findings may be of important translational value in indicating glutamatergic and neuroinflammatory-associated mechanisms as treatment targets, the currently applied ^1^H-MRS methodology is unlikely to have the required accuracy to predict response or pre-select patients for clinical trials at an individual level. This may be facilitated in the future through further advances in ^1^H-MRS methodology, including the use of higher-field strengths, which can provide greater spectral resolution and improved signal-to-noise ratio, as well as through integration of ^1^H-MRS measures with additional biomarkers and consideration of more nuanced and temporal patterns of clinical response.

## Conclusion

This large-scale mega-analysis provides the most robust evidence to date that elevated glutamatergic and inflammatory-related metabolites (glutamate, Glx, choline, mI) in the medial frontal cortex are associated with a poor response to antipsychotic medication in psychosis. Our findings show that elevated Glx may predict non-response prospectively, while mI is a marker of treatment-resistance. Collectively, these results support a shift in therapeutic strategy for non-responsive patients, moving towards novel mechanisms that target glutamatergic and neuroinflammatory pathways.

## Data Availability

The datasets used in this mega-analysis were obtained from previously published studies and were shared with the authors under data sharing agreements. These data are not publicly available because they are owned by the original investigators. Researchers wishing to access the underlying datasets should contact the corresponding authors of the original studies.

## Funding and disclosures

## Conflicts of Interest

K.B.B received lecture fees from Lundbeck Pharma A/S. B.E. is part of the Advisory Board of Boehringer Ingelheim, Lundbeck Pharma A/S, and Orion Pharma A/S; and has received lecture fees from Boehringer Ingelheim, Otsuka Pharma Scandinavia AB, and Lundbeck Pharma A/S. F.R.M has served as a consultant for Boehringer Ingelheim. M.M. is an employee of MSD New Zealand and engaged in this research as a private individual, not as a company affiliate. As such the principles, ideas and perspectives provided are the authors’ own and not those of MSD New Zealand. S.N. has received research support, manuscript fees or speaker’s honoraria from Asahi Quality & Innovations, Ltd., Teijin Pharma, Sumitomo Pharma, Meiji Seika Pharma, Otsuka, PDR pharma, and MSD within the past three years. E.v.d.G has received research support from NWO, ZonMw, Hersenstichting, Alzheimer Nederland, Health∼Holland and KWF. E.v.d.G has conducted contract research for Heuron Inc., AC Immune and Roche, and holds a consultancy agreement with IXICO for analysing PET scans. All support was paid to the institution. J.T.R.W. has received grant funding from Takeda and Akrivia Health for work unrelated to the current research. A.E. has received consultancy fees from Leal Therapeutics.

## Funding

B.K. is supported by a UK Medical Research Council PhD studentship (MR/N013700/1). K.B.B received a Ph.D grant from the Faculty of Health and Medical Sciences, University of Copenhagen for the data included in current study. A.d.B. has received research support from Janssen Italia, Lundbeck I, and OtsukaItalia and lecture fees for unrestricted educational meeting from Chiesi, Lundbeck Italian, Roche, Sunovion, Viatris, HIKMA, Tabuk, Recordati, Angelini, Gedeon-Richter and Takeda; he has served on advisory boards for Eli Lilly, Jansen, Lundbeck, Otsuka, Roche, Takeda, Chiesi, Recordati, Angelini, Viatris, Newron, Gedeon Richter. C.d.l.F.S. was supported by Secretaría de Ciencia, Humanidades, Tecnología e Innovación (SECIHTI), Mexico, Grants No. 261895 and 430. C.d.l.F.S. and F.R.M were supported by SECIHTI’s Sistema Nacional de Investigadoras e Investigadores. B.Y.J. has been the leader of a Lundbeck Foundation Centre of Excellence for Clinical Intervention and Neuropsychiatric Schizophrenia Research (CINS) (January 2009 – December 2021), which was partially financed by an independent grant from the Lundbeck Foundation based on international review and partially financed by the Mental Health Services in the Capital Region of Denmark, the University of Copenhagen, and other foundations. All grants are the property of the Mental Health Services in the Capital Region of Denmark and administrated by them. S.N. has received grants from Japan Society for the Promotion of Science (18H02755, 22H03002), Japan Agency for Medical Research and development (AMED: JP24wm0625302, JP24wm0625307), Japan Research Foundation for Clinical Pharmacology, Naito Foundation, Takeda Science Foundation, Watanabe Foundation, Osakeno-Kagaku Foundation, and Astellas Foundation within the past three years. S.M.L has been paid by Kynexis and Wellcome for educational sessions with their employees. J.T.R.W. is funded by a UKRI MRC Programme Grant (MR/Y004094/1). This paper represents independent research part-funded by the National Institute for Health and Care Research (NIHR) Biomedical Research Centre at South London and Maudsley NHS Foundation Trust and King’s College London. The views expressed are those of the authors and not necessarily those of the NHS, the NIHR or the Department of Health and Social Care. For the purposes of open access, the author has applied a CC BY public copyright licence to any Author Accepted Manuscript version arising from this submission.

## Notes

### Clinical Protocols

https://www.crd.york.ac.uk/PROSPERO/view/CRD42022346139

### Funding Statement

B.K. is supported by a UK Medical Research Council PhD studentship (MR/N013700/1). K.B.B received a PhD grant from the Faculty of Health and Medical Sciences, University of Copenhagen for the data included in the current study. A.d.B. has received research support from Janssen Italia, Lundbeck I, and Otsuka Italia, and lecture fees for unrestricted educational meetings from Chiesi, Lundbeck Italian, Roche, Sunovion, Viatris, HIKMA, Tabuk, Recordati, Angelini, Gedeon-Richter and Takeda; he has served on advisory boards for Eli Lilly, Jansen, Lundbeck, Otsuka, Roche, Takeda, Chiesi, Recordati, Angelini, Viatris, Newron, Gedeon Richter. C.d.l.F.S. was supported by Secretaria de Ciencia, Humanidades, Tecnologia e Innovacion (SECIHTI), Mexico, Grants No. 261895 and 430. C.d.l.F.S. and F.R.M. were supported by SECIHTI's Sistema Nacional de Investigadoras e Investigadores. B.Y.J. has been the leader of a Lundbeck Foundation Centre of Excellence for Clinical Intervention and Neuropsychiatric Schizophrenia Research (CINS) (January 2009 - December 2021), which was partially financed by an independent grant from the Lundbeck Foundation based on international review and partially financed by the Mental Health Services in the Capital Region of Denmark, the University of Copenhagen, and other foundations. All grants are the property of the Mental Health Services in the Capital Region of Denmark and administered by them. S.N. has received grants from the Japan Society for the Promotion of Science (18H02755, 22H03002), the Japan Agency for Medical Research and Development (AMED: JP24wm0625302, JP24wm0625307), the Japan Research Foundation for Clinical Pharmacology, the Naito Foundation, the Takeda Science Foundation, the Watanabe Foundation, the Osakeno-Kagaku Foundation, and the Astellas Foundation within the past three years. J.T.R.W. is funded by a UKRI MRC Programme Grant (MR/Y004094/1). This paper represents independent research part-funded by the National Institute for Health and Care Research (NIHR) Biomedical Research Centre at South London and Maudsley NHS Foundation Trust and King's College London. The views expressed are those of the authors and not necessarily those of the NHS, the NIHR or the Department of Health and Social Care.

### Author Declarations

The current study is a mega-analysis of separate, previously collected datasets. All data were fully anonymised before being shared with the study team under data sharing agreements. Each contributing study received ethical approval from the respective Ethics Committees or Institutional Review Boards of their host institutions prior to data collection. As this research involved secondary analysis of anonymised data, no additional ethical approval was required.

