## Supplementary File for "^1^H-MRS metabolites in antipsychotic-responsive versus non-responsive psychosis: a meta- and mega-analysis"

### Supplementary materials

**Supplementary Table of Contents**

**Supplementary Figure 1.** PRISMA diagram ........................................................... **p. 3**

**Supplementary Table 1.** Studies that contributed individual patient data ............... **pp. 4–6**

**Supplementary Table 2.** Response and non-response criteria of included studies… **pp. 7-8**

**Supplementary Table 3.** Mega-analysis dataset ...................................................... **p. 9**

**Supplementary Table 4.** Homoscedastic vs. Heteroscedastic models .................... **pp. 10-11**

**Supplementary Figure 2.** MFC voxel placements ……............................................ **p. 12**

**Supplementary Table 5.** Log-transformed linear mixed models…………………. **p.13**

**Supplementary Table 6.** Estimated marginal means of $^{1}$H-MRS metabolites in prospective first-episode psychosis studies ................................................................................... **pp. 14-15**

**Supplementary Table 7.** Estimated marginal means of $^{1}$H-MRS metabolites including treatment-resistant samples ......................................................................................... **pp. 16-17**

**Sensitivity Analysis.** CSF-corrected metabolites ................................................... **p. 18**

**Supplementary Table 8.** Estimated marginal means of $^{1}$H-MRS metabolites with CSF correction ......................................................................................................... **pp. 19-20**

**Meta-analysis Methods.** Literature search, extraction, and statistical analyses ...... **pp. 21-22**

**Supplementary Table 9.** Studies included in meta-analyses.................................. **pp. 23-27**

**Supplementary Table 10.** Results of meta-analysis of mean differences ................ **pp. 28-29**

**Supplementary Figure 3.** Forest plot for meta-analysis of mean differences .......... **p. 30**

**Supplementary Table 11.** Results of meta-analysis of variability........................... **pp. 31-32**

**Supplementary Figure 4.** Forest plot for meta-analysis of variability ..................... **p. 33**

**References** ........................................................................................................ **pp. 34–36**

**Supplementary Figure 1.**

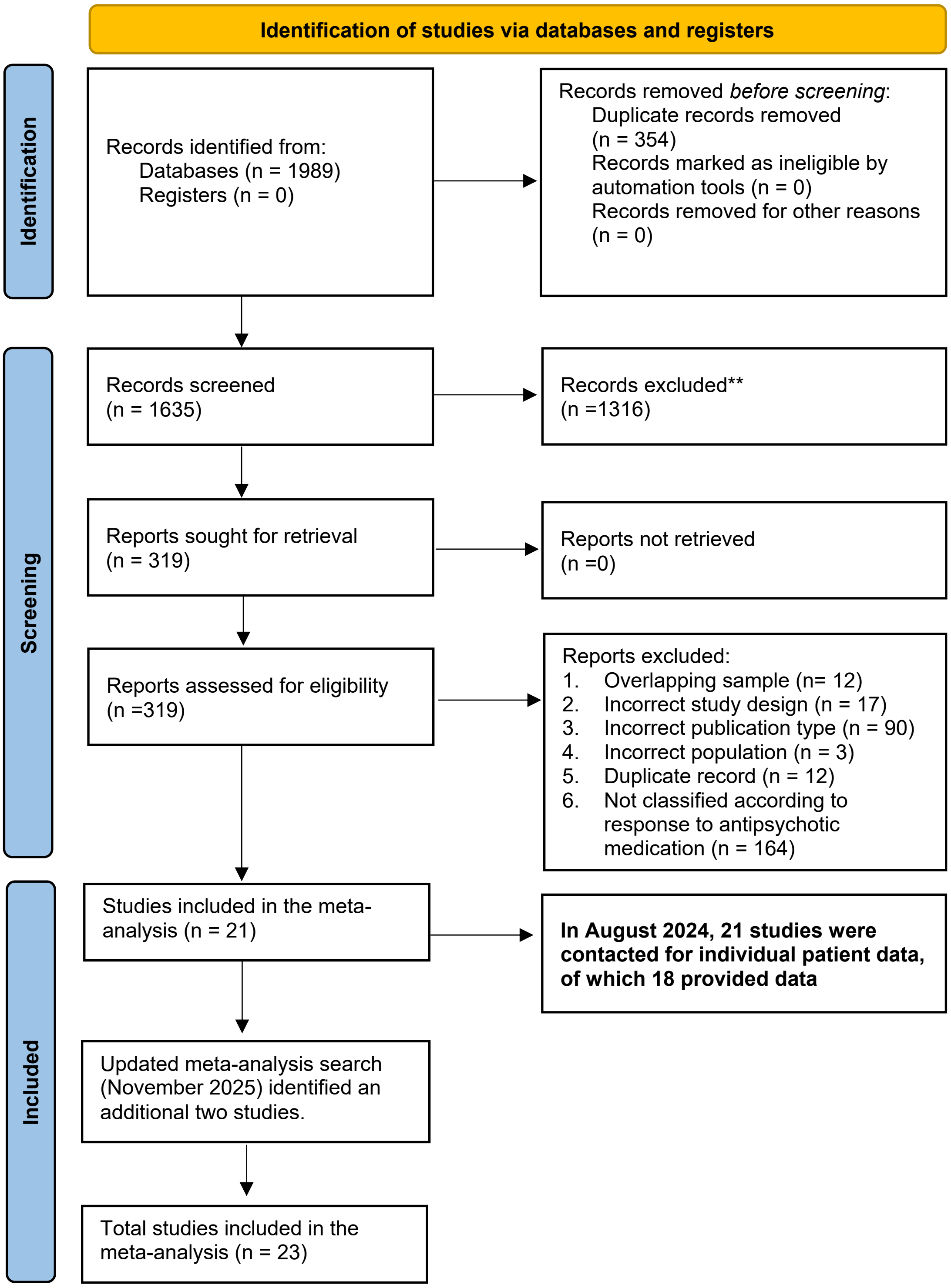

**Supplementary Table 1. Studies that contributed individual patient data**

| | | N participants | | | Metabolite data by brain region | | | | | Clinical and demographic data | | | | Subgroup analysis | | |
|---|---|---|---|---|---|---|---|---|---|---|---|---|---|---|---|---|
| Study (ref) | Year | R | NR | HC | Scanner tesla | MFC | DLPFC | Thalamus | BG | Age | Sex | PANSS | CPZ | Design | TRS sample | Correction method |
| Egerton (1) | 2012 | 15 | 17 | 0 | 3T | Glu, Glx, NAA, Cho, mI | ✗ | Glu, Glx, NAA, Cho, mI | ✗ | ✓ | ✓ | ✓ | ✓ | Cross-sectional | ✗ | Cr |
| McIlwain (2) | 2015 | 18 | 30 | 18 | 3T | Glu, Glx, NAA, Cho | Glu, Glx, NAA, Cho | ✗ | Glu, Glx, NAA, Cho | ✓ | ✓ | ✓ | ✓ | Cross-sectional | ✓ | CSF |
| Mouchlianitis (3) | 2015 | 19 | 18 | 0 | 3T | Glu | ✗ | ✗ | ✗ | ✓ | ✓ | ✓ | ✓ | Cross-sectional | ✓ | Cr |
| Egerton (Site A) (4) | 2018 | 17 | 5 | 0 | 3T | Glu, Glx, NAA, Cho, mI | ✗ | Glu, Glx, NAA, Cho, mI | ✗ | ✓ | ✓ | ✓ | ✓ | Prospective | ✗ | Cr |
| Egerton (Site B) (4) | | 13 | 13 | 0 | 3T | Glu, Glx, NAA, Cho, mI | ✗ | Glu, Glx, NAA, Cho, mI | ✗ | ✓ | ✓ | ✓ | ✓ | Prospective | ✗ | Cr |
| Egerton (Site C) (4) | | 8 | 6 | 0 | 3T | Glu, Glx, NAA, Cho, mI | ✗ | Glu, Glx, NAA, Cho, mI | ✗ | ✓ | ✓ | ✓ | ✓ | Prospective | ✗ | Cr |
| Iwata (5) | 2019 | 21 | 53 | 26 | 3T | Glu, Glx, NAA, Cho, mI | Glu, Glx, NAA, Cho, mI | ✗ | Glu, Glx, NAA, Cho, mI | ✓ | ✓ | ✓ | ✓ | Cross-sectional | ✓ | CSF and Cr |
| Bojesen (6) | 2020 | 11 | 21 | 35 | 3T | Glu, Glx, NAA, Cho, mI | ✗ | Glu, Glx, NAA, Cho, mI | ✗ | ✓ | ✓ | ✓ | ✓ | Prospective | ✗ | CSF and Cr |
| Dempster (7) | 2020 | 9 | 17 | 27 | 7T | Glu, Glx and GSH | ✗ | ✗ | ✗ | ✓ | ✓ | ✗ | ✗ | Prospective | ✗ | CSF and Cr |
| Egerton (Site A) (8) | 2021 | 16 | 18 | 0 | 3T | Glu, Glx, NAA, Cho, mI | ✗ | Glu, Glx, NAA, Cho, mI | Glu, Glx, NAA, Cho, mI | ✓ | ✓ | ✓ | ✓ | Cross-sectional | ✗ | CSF and Cr |

| | | | | | | | | | | | | | | | | |
|---|---|---|---|---|---|---|---|---|---|---|---|---|---|---|---|---|
| Egerton (Site B) (8) | | 17 | 16 | 0 | 3T | Glu, Glx, NAA, Cho, mI | ✗ | Glu, Glx, NAA, Cho, mI | Glu, Glx, NAA, Cho, mI | ✓ | ✓ | ✓ | ✓ | Cross-sectional | ✗ | CSF and Cr |
| Egerton (Site C) (8) | 2021 | 8 | 5 | 0 | 3T | Glu, Glx, NAA, Cho, mI | ✗ | Glu, Glx, NAA, Cho, mI | Glu, Glx, NAA, Cho, mI | ✓ | ✓ | ✓ | ✓ | Cross-sectional | ✗ | CSF and Cr |
| Egerton (Site D) (8) | | 7 | 8 | 0 | 3T | Glu, Glx, NAA, Cho, mI | ✗ | Glu, Glx, NAA, Cho, mI | Glu, Glx, NAA, Cho, mI | ✓ | ✓ | ✓ | ✓ | Cross-sectional | ✗ | CSF and Cr |
| Horne (9) | 2021 | 21 | 19 | 20 | 3T | Glu, Glx, NAA, Cho, mI | ✗ | ✗ | ✗ | ✓ | ✓ | ✓ | ✓ | Cross-sectional | ✓ | Cr |
| Iwata (10) | 2021 | 21 | 51 | 26 | 3T | GSH | ✗ | ✗ | ✗ | ✓ | ✓ | ✓ | ✓ | Cross-sectional | ✓ | CSF |
| Yang (11) | 2021 | 29 | 14 | 0 | 7T | Glu, Glx, NAA, GABA, GSH | Glu, Glx, NAA, GABA, GSH | Glu, Glx, NAA, GABA, GSH | ✗ | ✓ | ✓ | ✗ | ✓ | Prospective | ✓ | CSF and Cr |
| Huang (12) | 2022 | 34 | 37 | 19 | 1.5T | Glu, Glx | ✗ | ✗ | ✗ | ✓ | ✓ | ✓ | ✓ | Cross-sectional | ✓ | NAA ratio |
| Matrone (13) | 2022 | 0 | 20 | 20 | 1.5T | Glu, Glx | ✗ | ✗ | ✗ | ✓ | ✓ | ✓ | ✓ | Cross-sectional | ✓ | Cr |
| Reyes-Madrigal (14) | 2022 | 29 | 19 | 0 | 3T | ✗ | ✗ | ✗ | Glu, Glx, NAA, mI | ✓ | ✓ | ✓ | ✓ | Prospective | ✗ | CSF |
| Ueno (15) | 2022 | 16 | 47 | 35 | 3T | GABA | ✗ | ✗ | ✗ | ✓ | ✓ | ✓ | ✓ | Cross-sectional | ✓ | CSF and Cr |
| Egerton (Site A) (16) | 2023 | 20 | 10 | 0 | 3T | Glu, Glx, NAA, Cho, mI | ✗ | ✗ | Glu, Glx, NAA, Cho, mI | ✓ | ✓ | ✓ | ✓ | Prospective | ✗ | CSF and Cr |
| Egerton (Site B) (16) | | 8 | 8 | 0 | 3T | Glu, Glx, NAA, Cho, mI | ✗ | ✗ | Glu, Glx, NAA, Cho, mI | ✓ | ✓ | ✓ | ✓ | Prospective | ✗ | CSF and Cr |
| Fan (17) | 2023 | 27 | 15 | 41 | 3T | Glu, Glx, NAA, Cho, mI | ✗ | ✗ | ✗ | ✓ | ✓ | ✓ | ✗ | Prospective | ✗ | CSF |
| Van Der Pluijm (18) | 2024 | 40 | 10 | 19 | 3T | Glu, Glx, NAA, Cho, mI, GABA | ✗ | ✗ | ✗ | ✓ | ✓ | ✓ | ✓ | Prospective | ✓ | CSF and Cr |

Note: Where studies acquired data at more than one site data are treated separately. If a study contributed data with more than one correction method, CSF corrected metabolites were prioritised over Cr scaled metabolites. Abbreviations: Cho: choline; Cr: Creatine; DLPFC: dorsolateral prefrontal cortex; GABA: gamma-aminobutyric acid; Glu: glutamate; Glx: glutamate+glutamine; GSH: glutathione; HC: healthy controls; MFC: medial frontal cortex; mI: myo-inositol; NAA: N-acetylaspartate; NR: non-responder group; R: responder group.

**Supplementary Table 2. Definitions of treatment response and non-response across included studies**

| Study | Year | Definition of treatment response | Definition of non-response / treatment resistance |
|---|---|---|---|
| Egerton | 2012 | Symptomatic remission based on Andreasen criteria (19): scores of ≤3 (mild) on PANSS items: P1, P2, P3, N1, N4, N6, G5, G9. | Scores ≥4 (moderate) on any of the same PANSS items. |
| McIlwain | 2015 | First-line responders taking a second-generation (non-clozapine) antipsychotic, with mild illness or better on CGI. | Mild illness or better after clozapine monotherapy (TRS); combined antipsychotics after clozapine monotherapy failed (UTRS). |
| Mouchlianitis | 2015 | Symptomatic remission: no PANSS item ≤3, never had clozapine, no relapse in prior 6 months. | TRS defined using modified Kane criteria (20) and no current or previous clozapine use. |
| Egerton | 2018 | Andreasen remission criteria (19), excluding the 6-month requirement. | Patients not meeting response criteria. |
| Iwata | 2019 | [1] CGI-Severity ≤3; [2] all PANSS positive items ≤3; [3] no relapse in prior 3 months. | TRS defined using TRIPP criteria (21).<br>UTRS: non-response to ≥6 weeks clozapine at ≥300 mg/day. |
| Bojesen | 2020 | Andreasen remission criteria (19), excluding the 6-month requirement. | Patients not meeting response criteria. |
| Dempster | 2020 | Andreasen remission criteria (19):, excluding the 6-month requirement. | Patients not meeting response criteria. |
| Egerton | 2021 | [1] treatment with only 1 antipsychotic since onset (unless changed for side effects); [2] CGI-SCH <4; [3] PANSS total <60 (22); and [4] CRS >3 (23) | Non-responders met all: [1] ≥2 antipsychotic trials >4 weeks at therapeutic doses; [2] CGI-SCH >3; [3] PANSS ≥70 and [4] CRS >3. |

| | | | |
|---|---|---|---|
| Horne | 2021 | Symptomatic remission: PANSS items ≤3(20), stable for ≥6 months; stable antipsychotic dose for ≥6 months (19). | Persistent symptoms (≥4 on ≥2 PANSS positive items), non-response to ≥2 antipsychotic trials of 4–6 weeks, illness duration ≥5 years with no good functional period. |
| Iwata | 2021 | As per Iwata et al. (2019). | As per Iwata et al. (2019). |
| Yang | 2021 | Patients not meeting TRS criteria. | TRS: ≥2 non-clozapine antipsychotics previously used and/or currently taking clozapine. |
| Huang | 2022 | Patients not meeting TRS criteria. | TRS defined using TRIPP criteria (21). |
| Matrone | 2022 | Not applicable (no responder group). | TRS defined using TRIPP criteria (21). |
| Reyes-Madrigal | 2022 | ≥40% reduction in PANSS positive subscale. | <40% reduction in PANSS positive subscale. |
| Ueno | 2022 | As per Iwata et al. (2019). | As per Iwata et al. (2019). |
| Egerton | 2023 | >20% reduction in PANSS total score from baseline to 6 weeks. | <20% reduction in PANSS total score at 6 weeks. |
| Fan | 2023 | >50% reduction in PANSS total score from baseline to follow-up. | <50% reduction in PANSS total score from baseline to follow-up. |
| Van der Pluijm | 2024 | Patients not meeting TR criteria. | TR: non-response to ≥2 antipsychotic trials plus ≥1 PANSS item (P1, P2, P3, G5, G9) rated ≥ 4 or currently taking clozapine. |

Abbreviations: CGI-SCH: clinical global impression - schizophrenia scale; CRS: compliance rating scale; non-TRS: non treatment resistant schizophrenia; PANSS: positive and negative syndrome scale; TRIPP: treatment response and resistance in psychosis working group consensus; TRS: treatment-resistant schizophrenia; UTRS: ultra-treatment-resistant schizophrenia.

**Supplementary Table 3.** Mega-analysis dataset

| | NR | R | HC | P value |
|---|---|---|---|---|
| Sex M/F | 299/128 | 340/136 | 170/89 | P = 0.258 |
| Age | 32.96 (12.44) | 30.36 (11.48) | 30.96 (12.44) | P = 0.002* |
| CPZ dose | 496.39 (302.95) | 345.55 (222.38) | -- | P <.001 |
| PANSS total | 70.90 (18.02) | 51.13 (12.16) | -- | P<.001 |

Note: Data are presented as Mean (SD) unless otherwise stated. Group differences were established using chi-squared for categorical variables (sex) and ANOVA (age) or two-sample-tests for continuous variables (CPZ and PANSS total scores). Abbreviations: HC: healthy controls; NR: non-responder group; R: responder group.

**Supplementary Table 4.** Statistical comparison of homoscedastic vs heteroscedastic model fit

| Metabolite | Region | Model | DF | AIC | BIC | logLik | L Ratio | P value |
|---|---|---|---|---|---|---|---|---|
| **Glu** | | | | | | | | |
| | MFC | 1 | 7 | 3553.56 | 3586.99 | -1769.78 | | |
| | | 2 | 9 | 3513.40 | 3556.38 | -1747.70 | 44.17 | **<.001** |
| | DLPFC | 1 | 7 | 618.83 | 641.45 | -302.42 | | |
| | | 2 | 9 | 612.92 | 642.00 | -297.46 | 9.91 | **0.007** |
| | Thalamus | 1 | 7 | 1287.35 | 1312.08 | -636.67 | | |
| | | 2 | 9 | 949.78 | 981.58 | -465.89 | 341.57 | **<.001** |
| | Basal ganglia | 1 | 7 | 3531.72 | 3557.88 | -1758.86 | | |
| | | 2 | 9 | 2782.34 | 2815.97 | -1382.17 | 753.38 | **<.001** |
| **Glx** | | | | | | | | |
| | MFC | 1 | 7 | 3194.30 | 3226.27 | -1590.15 | | |
| | | 2 | 9 | 3185.89 | 3226.99 | -1583.94 | 12.41 | **0.002** |
| | Thalamus | 1 | 7 | 1305.39 | 1329.02 | -645.70 | | |
| | | 2 | 9 | 1042.75 | 1073.12 | -512.37 | 266.65 | **<.001** |
| | Basal ganglia | 1 | 7 | 3778.28 | 3804.46 | -1882.14 | | |
| | | 2 | 9 | 3126.95 | 3160.61 | -1554.47 | 655.33 | **<.001** |
| **NAA** | | | | | | | | |
| | MFC | 1 | 7 | 2346.22 | 2378.04 | -1166.11 | | |
| | | 2 | 9 | 2328.01 | 2368.92 | -1155.00 | 22.21 | **<.001** |
| | DLPFC | 1 | 7 | 558.15 | 580.77 | -272.07 | | |
| | | 2 | 9 | 553.67 | 582.75 | -267.84 | 8.48 | **0.014** |
| | Thalamus | 1 | 7 | 1318.53 | 1343.49 | -652.27 | | |
| | | 2 | 9 | 919.15 | 951.23 | -450.57 | 403.39 | **<.001** |
| | Basal ganglia | 1 | 7 | 3336.59 | 3362.79 | -1661.30 | | |
| | | 2 | 9 | 2224.91 | 2258.59 | -1103.45 | 1115.69 | **<.001** |
| **mI** | | | | | | | | |
| | MFC | 1 | 5 | 1912.94 | 1934.96 | -951.47 | | |
| | | 2 | 7 | 1907.61 | 1938.45 | -946.81 | 9.32 | **0.009** |
| | Thalamus | 1 | 5 | 741.35 | 758.27 | -365.67 | | |
| | | 2 | 7 | 561.27 | 584.96 | -273.64 | 184.07 | **<.001** |
| | Basal ganglia | 1 | 5 | 2866.84 | 2884.83 | -1428.42 | | |
| | | 2 | 7 | 2555.04 | 2580.22 | -1270.52 | 315.80 | **<.001** |
| **Cho** | | | | | | | | |
| | MFC | 1 | 7 | 552.54 | 583.92 | -269.27 | | |
| | | 2 | 9 | 528.81 | 569.16 | -255.41 | 27.73 | **<.001** |
| | Thalamus | 1 | 7 | 488.21 | 512.00 | -237.11 | | |
| | | 2 | 9 | 166.11 | 196.70 | -74.06 | 326.10 | **<.001** |

| | | | | | | | | |
|---|---|---|---|---|---|---|---|---|
| | Basal ganglia | 1 | 7 | 2467.31 | 2492.45 | -1226.65 | | |
| | | 2 | 9 | 1967.62 | 1999.93 | -974.81 | 503.69 | **<.001** |
| **GABA** | | | | | | | | |
| | MFC | 1 | 5 | 183.58 | 201.25 | -86.79 | | |
| | | 2 | 7 | 187.27 | 212.00 | -86.63 | 0.31 | 0.855 |
| **GSH** | | | | | | | | |
| | MFC | 1 | 5 | 51.72 | 67.83 | -20.86 | | |
| | | 2 | 7 | 22.71 | 45.26 | -4.36 | 33.01 | **<.001** |

Note: Homoscedastic (Model 1) and heteroscedastic (Model 2) linear mixed models were compared, with the best-fitting model selected based on AIC and likelihood ratio tests (LRT). Significant results are indicated in bold. Abbreviations: Cho: choline; DLPFC: dorsolateral prefrontal cortex; GABA: gamma-aminobutyric acid; Glu: glutamate; Glx: glutamate+glutamine; GSH: glutathione; MFC: medial frontal cortex; mI: myo-inositol; NAA: N-acetylaspartate.

**Supplementary Figure 2.**

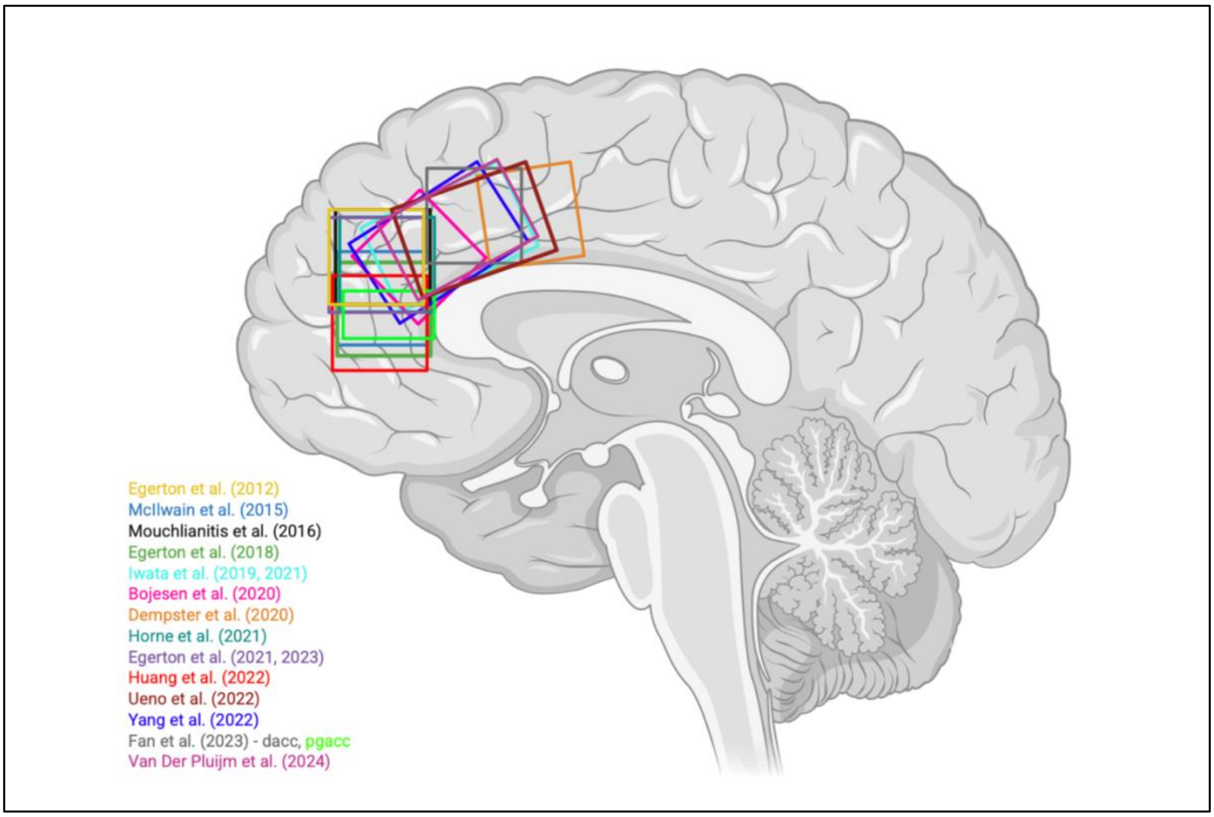


Illustration of MFC $^1$H-MRS voxel placements for studies included in the mega-analysis. Created in BioRender. King, B. (2026) https://BioRender.com/jb829b4

**Supplementary Table 5.** Log-transformed linear mixed models - $^{1}$H-MRS metabolite group effects

| Metabolite | Fixed effect | N cohorts | N participants | df | F | P |
|---|---|---|---|---|---|---|
| **Glu** | | | | | | |
| DLPFC | Group (NR, R, HC) | 3 | 187 | 180 | 6.20 | 0.003 |
| Basal Ganglia | Group (NR, R, HC) | 9 | 310 | 297 | 0.16 | 0.855 |
| **Glx** | | | | | | |
| Basal Ganglia | Group (NR, R, HC) | 9 | 311 | 298 | 0.49 | 0.612 |
| **NAA** | | | | | | |
| DLPFC | Group (NR, R, HC) | 3 | 187 | 180 | 0.98 | 0.379 |
| Basal Ganglia | Group (NR, R, HC) | 9 | 312 | 298 | 0.15 | 0.859 |
| **Cho** | | | | | | |
| Basal Ganglia | Group (NR, R, HC) | 8 | 268 | 255 | 0.15 | 0.857 |
| **mI** | | | | | | |
| Basal Ganglia | Group (NR, R, HC) | 8 | 270 | 258 | 0.48 | 0.622 |

Abbreviations: Cho: choline; DLPFC: dorsolateral prefrontal cortex; Glu: glutamate; Glx: glutamate+glutamine; HC: healthy controls; mI: myo-inositol; NAA: N-acetylaspartate; NR: non-responder group; R: responder group.

**Supplementary Table 6.** Estimated marginal means (EMMs) of $^{1}$H-MRS metabolites by group in prospective first-episode psychosis studies.

| | Group | N | EMM | SE | Lower CI | Upper CI |
|---|---|---|---|---|---|---|
| **Glu** | | | | | | |
| MFC | Non-Responder | 114 | 8.96 | 2.05 | 4.31 | 13.60 |
| | Responder | 181 | 8.48 | 2.04 | 3.86 | 13.10 |
| | HC | 95 | 8.69 | 2.05 | 4.05 | 13.33 |
| Thalamus | Non-Responder | 50 | 3.32 | 1.27 | -0.22 | 6.85 |
| | Responder | 76 | 3.38 | 1.27 | -0.15 | 6.91 |
| | HC | 31 | 3.12 | 1.28 | -0.44 | 6.67 |
| Basal Ganglia | Non-Responder | 35 | 11.09 | 1.13 | 6.23 | 15.96 |
| | Responder | 46 | 10.92 | 1.12 | 6.09 | 15.74 |
| **Glx** | | | | | | |
| MFC | Non-Responder | 100 | 12.39 | 3.01 | 5.46 | 19.33 |
| | Responder | 153 | 11.57 | 3.01 | 4.63 | 18.50 |
| | HC | 95 | 11.81 | 3.01 | 4.86 | 18.76 |
| Thalamus | Non-Responder | 38 | 3.35 | 1.88 | -2.62 | 9.32 |
| | Responder | 51 | 3.48 | 1.87 | -2.49 | 9.44 |
| | HC | 32 | 3.59 | 1.89 | -2.43 | 9.61 |
| Basal Ganglia | Non-Responder | 35 | 15.19 | 1.00 | 10.88 | 19.50 |
| | Responder | 46 | 14.78 | 0.98 | 10.56 | 18.99 |
| **NAA** | | | | | | |
| MFC | Non-Responder | 97 | 7.78 | 1.84 | 3.53 | 12.04 |
| | Responder | 171 | 7.40 | 1.84 | 3.15 | 11.65 |
| | HC | 95 | 7.61 | 1.85 | 3.35 | 11.87 |
| Thalamus | Non-Responder | 52 | 3.79 | 1.28 | 0.23 | 7.35 |
| | Responder | 79 | 3.89 | 1.28 | 0.33 | 7.44 |
| | HC | 32 | 4.08 | 1.29 | 0.51 | 7.65 |
| Basal Ganglia | Non-Responder | 35 | 9.99 | 0.96 | 5.84 | 14.14 |
| | Responder | 46 | 10.00 | 0.96 | 5.88 | 14.12 |
| **Cho** | | | | | | |
| MFC | Non-Responder | 83 | 1.62 | 0.43 | 0.60 | 2.64 |
| | Responder | 144 | 1.56 | 0.43 | 0.55 | 2.57 |
| | HC | 95 | 1.46 | 0.43 | 0.44 | 2.48 |
| Thalamus | Non-Responder | 39 | 0.60 | 0.31 | -0.38 | 1.58 |
| | Responder | 52 | 0.63 | 0.31 | -0.34 | 1.61 |
| | HC | 32 | 0.70 | 0.31 | -0.28 | 1.68 |
| Basal Ganglia | Non-Responder | 16 | 1.12 | 0.52 | -5.54 | 7.78 |
| | Responder | 21 | 0.97 | 0.51 | -5.55 | 7.49 |
| **mI** | | | | | | |
| MFC | Non-Responder | 83 | 4.90 | 1.28 | 1.87 | 7.93 |

| | | | | | | |
|---|---|---|---|---|---|---|
| | Responder | 144 | 4.77 | 1.28 | 1.74 | 7.79 |
| | HC | 95 | 4.68 | 1.28 | 1.65 | 7.71 |
| Thalamus | Non-Responder | 38 | 1.34 | 0.65 | -0.72 | 3.40 |
| | Responder | 51 | 1.31 | 0.65 | -0.75 | 3.36 |
| | HC | 32 | 1.55 | 0.65 | -0.53 | 3.63 |
| Basal Ganglia | Non-Responder | 35 | 4.48 | 0.75 | 1.25 | 7.71 |
| | Responder | 46 | 4.72 | 0.83 | 1.17 | 8.28 |

Abbreviations: Cho: choline; CI: confidence interval; DLPFC: dorsolateral prefrontal cortex; EMMs: estimated marginal means; GABA: gamma-aminobutyric acid; Glu: glutamate; Glx: glutamate + glutamine; GSH: glutathione; HC: healthy controls; mI: myo-inositol; MFC: medial frontal cortex; NAA: N-acetylaspartate; SE: standard error.

**Supplementary Table 7.** Estimated marginal means (EMMs) of $^{1}$H-MRS metabolites by group, including treatment-resistant.

| Metabolite | Group | N | EMM | SE | Lower CI | Upper CI |
|---|---|---|---|---|---|---|
| **Glu** | | | | | | |
| MFC | Treatment Resistant | 174 | 8.27 | 1.51 | 5.10 | 11.43 |
| | Non-Responder | 151 | 8.61 | 1.52 | 5.42 | 11.79 |
| | Responder | 352 | 8.13 | 1.51 | 4.97 | 11.29 |
| | HC | 173 | 8.03 | 1.51 | 4.87 | 11.20 |
| Thalamus | Treatment Resistant | 13 | 4.21 | 1.25 | 1.25 | 7.18 |
| | Non-Responder | 87 | 4.92 | 1.33 | 1.78 | 8.07 |
| | Responder | 122 | 4.33 | 1.23 | 1.42 | 7.24 |
| | HC | 31 | 4.05 | 1.25 | 1.10 | 7.01 |
| Basal Ganglia | Treatment Resistant | 66 | 9.20 | 1.62 | 5.48 | 12.93 |
| | Non-Responder | 80 | 25.11 | 15.82 | -11.37 | 61.59 |
| | Responder | 126 | 9.94 | 1.43 | 6.63 | 13.25 |
| | HC | 38 | 9.01 | 1.63 | 5.25 | 12.78 |
| **Glx** | | | | | | |
| MFC | Treatment Resistant | 123 | 12.49 | 2.23 | 7.76 | 17.21 |
| | Non-Responder | 151 | 12.78 | 2.23 | 8.06 | 17.50 |
| | Responder | 284 | 12.08 | 2.22 | 7.37 | 16.78 |
| | HC | 153 | 12.01 | 2.22 | 7.29 | 16.72 |
| Basal Ganglia | Treatment Resistant | 67 | 13.43 | 3.22 | 5.99 | 20.86 |
| | Non-Responder | 80 | 36.57 | 23.10 | -16.70 | 89.85 |
| | Responder | 126 | 15.05 | 2.79 | 8.61 | 21.49 |
| | HC | 38 | 12.94 | 3.23 | 5.50 | 20.39 |
| **NAA** | | | | | | |
| MFC | Treatment Resistant | 119 | 8.22 | 1.39 | 5.26 | 11.18 |
| | Non-Responder | 134 | 8.24 | 1.39 | 5.28 | 11.20 |
| | Responder | 289 | 7.92 | 1.39 | 4.97 | 10.87 |
| | HC | 154 | 7.89 | 1.39 | 4.93 | 10.84 |
| Thalamus | Treatment Resistant | 13 | 5.10 | 1.46 | 1.66 | 8.54 |
| | Non-Responder | 90 | 5.69 | 1.51 | 2.12 | 9.26 |
| | Responder | 126 | 5.26 | 1.43 | 1.88 | 8.64 |
| | HC | 32 | 5.48 | 1.44 | 2.08 | 8.88 |
| Basal Ganglia | Treatment Resistant | 67 | 8.12 | 1.09 | 5.61 | 10.63 |
| | Non-Responder | 80 | 19.35 | 11.24 | -6.58 | 45.28 |
| | Responder | 126 | 8.22 | 1.06 | 5.77 | 10.68 |
| | HC | 39 | 8.01 | 1.08 | 5.52 | 10.50 |
| **Cho** | | | | | | |
| MFC | Treatment Resistant | 104 | 1.78 | 0.31 | 1.11 | 2.44 |
| | Non-Responder | 134 | 1.72 | 0.31 | 1.05 | 2.38 |
| | Responder | 262 | 1.67 | 0.31 | 1.01 | 2.32 |

| | | | | | | |
|---|---|---|---|---|---|---|
| | HC | 154 | 1.58 | 0.31 | 0.92 | 2.24 |
| Basal Ganglia | Treatment Resistant | 67 | 1.83 | 0.90 | -0.29 | 3.95 |
| | Non-Responder | 61 | 8.06 | 6.37 | -7.00 | 23.13 |
| | Responder | 101 | 2.34 | 0.88 | 0.26 | 4.41 |
| | HC | 39 | 1.76 | 0.90 | -0.36 | 3.88 |
| **mI** | | | | | | |
| MFC | Treatment Resistant | 82 | 6.14 | 0.95 | 4.10 | 8.18 |
| | Non-Responder | 134 | 5.48 | 0.94 | 3.46 | 7.50 |
| | Responder | 248 | 5.35 | 0.94 | 3.34 | 7.36 |
| | HC | 141 | 5.15 | 0.94 | 3.13 | 7.17 |
| Basal Ganglia | Treatment Resistant | 50 | 6.02 | 0.29 | 5.34 | 6.71 |
| | Non-Responder | 80 | 14.32 | 9.72 | -8.65 | 37.29 |
| | Responder | 114 | 6.54 | 1.67 | 2.58 | 10.51 |
| | HC | 26 | 5.77 | 0.25 | 5.17 | 6.37 |
| **GSH** | | | | | | |
| MFC | Treatment Resistant | 61 | 1.22 | 0.21 | 0.30 | 2.15 |
| | Non-Responder | 17 | 1.41 | 0.24 | 0.38 | 2.43 |
| | Responder | 55 | 1.30 | 0.22 | 0.37 | 2.23 |
| | HC | 25 | 1.29 | 0.22 | 0.36 | 2.21 |

*Note*: Individuals in the non-responder group meeting or not meeting treatment resistant criteria were treated as separate groups within this analysis. Abbreviations: Cho: choline; CI: confidence interval; DLPFC: dorsolateral prefrontal cortex; EMMs: estimated marginal means; GABA: gamma-aminobutyric acid; Glu: glutamate; Glx: glutamate + glutamine; GSH: glutathione; HC: healthy controls; mI: myo-inositol; MFC: medial frontal cortex; NAA: N-acetylaspartate; SE: standard error.

**Follow up analysis: CSF-corrected metabolites**

When the mega-analysis was limited to studies employing CSF-correction, MFC Glu ($F$ = 4.817, df = 549, $P$ = 0.008), Glx ($F$ = 5.368, df = 508, $P$ = 0.005), Cho ($F$ = 11.814, df = 482, $P$ <.001) and mI ($F$ = 13.821, df = 482, $P$ <.001) showed significant effects of group.

Post hoc pairwise comparisons revealed that MFC Glu (Estimate (E) = 0.521, df = 549, $P$ = 0.004, Glass's $\Delta$ = 0.29), Glx (E = 0.806, df = 508, $P$ = 0.003, Glass's $\Delta$ = 0.32) and mI (E = 0.495, df = 434, $P$ = 0.001, Glass's $\Delta$ = 0.40) were significantly elevated in the non-responder compared to responder group. The non-responder group also showed significant elevations in Glu (E = 0.510, df = 549, $P$ = 0.011, Glass's $\Delta$ = 0.28), Glx (E = 0.815, df = 508, $P$ = 0.004, Glass's $\Delta$ = 0.32), Cho (E = 0.206, df = 482, $P$ <.001, Glass's $\Delta$ = 0.52) and mI (E = 0.795, df = 434, $P$ <.001, Glass's $\Delta$ = 0.65) compared to the controls. In addition, Cho (E = 0.117, df = 482, $P$ = 0.006, Glass's $\Delta$ = 0.29) and mI (E = 0.300, df = 434, $P$ = 0.047, Glass's $\Delta$ = 0.25) showed significant elevations in the responder group in comparison to controls.

**Supplementary Table 8.** Estimated marginal means (EMMs) of $^{1}$H-MRS metabolites with CSF correction

| | Group | N | EMM | SE | Lower CI | Upper CI |
|---|---|---|---|---|---|---|
| **Glu** | | | | | | |
| MFC | Non-Responder | 210 | 12.65 | 1.44 | 9.53 | 15.78 |
| | Responder | 222 | 12.13 | 1.43 | 9.01 | 15.26 |
| | HC | 134 | 12.14 | 1.44 | 9.01 | 15.28 |
| DLPFC | Non-Responder | 84 | 7.07 | 3.11 | -6.32 | 20.47 |
| | Responder | 62 | 7.69 | 3.11 | -5.71 | 21.08 |
| | HC | 41 | 7.22 | 3.12 | -6.19 | 20.62 |
| Thalamus | Non-Responder | 65 | 8.37 | 0.83 | 5.73 | 11.01 |
| | Responder | 71 | 7.67 | 0.48 | 6.15 | 9.19 |
| | HC | 31 | 7.35 | 0.53 | 5.68 | 9.03 |
| Basal Ganglia | Non-Responder | 146 | 18.12 | 8.75 | -2.05 | 38.29 |
| | Responder | 126 | 10.11 | 1.42 | 6.82 | 13.39 |
| | HC | 38 | 9.25 | 1.62 | 5.52 | 12.99 |
| **Glx** | | | | | | |
| MFC | Non-Responder | 196 | 17.31 | 2.03 | 12.85 | 21.77 |
| | Responder | 194 | 16.51 | 2.02 | 12.05 | 20.96 |
| | HC | 134 | 16.50 | 2.03 | 12.03 | 20.97 |
| Thalamus | Non-Responder | 52 | 12.81 | 1.50 | 6.36 | 19.26 |
| | Responder | 44 | 11.81 | 0.83 | 8.24 | 15.38 |
| | HC | 32 | 11.65 | 0.89 | 7.80 | 15.49 |
| Basal Ganglia | Non-Responder | 147 | 26.43 | 12.74 | -2.96 | 55.82 |
| | Responder | 126 | 15.40 | 2.78 | 8.99 | 21.81 |
| | HC | 38 | 13.46 | 3.21 | 6.06 | 20.86 |
| **NAA** | | | | | | |
| MFC | Non-Responder | 193 | 11.16 | 1.25 | 8.41 | 13.92 |
| | Responder | 212 | 10.74 | 1.25 | 7.98 | 13.50 |
| | HC | 134 | 10.74 | 1.26 | 7.98 | 13.50 |
| DLPFC | Non-Responder | 84 | 7.87 | 2.96 | -4.88 | 20.62 |
| | Responder | 61 | 8.09 | 2.96 | -4.66 | 20.84 |
| | HC | 42 | 7.55 | 2.96 | -5.21 | 20.31 |
| Thalamus | Non-Responder | 65 | 9.47 | 1.31 | 5.31 | 13.63 |
| | Responder | 71 | 8.92 | 1.12 | 5.37 | 12.47 |
| | HC | 32 | 9.13 | 1.13 | 5.53 | 12.72 |
| Basal Ganglia | Non-Responder | 147 | 14.24 | 6.19 | -0.04 | 28.52 |
| | Responder | 126 | 8.23 | 1.06 | 5.78 | 10.68 |
| | HC | 39 | 8.02 | 1.08 | 5.53 | 10.51 |
| **Choline** | | | | | | |
| MFC | Non-Responder | 178 | 2.40 | 0.27 | 1.79 | 3.01 |

| | | | | | | |
|---|---|---|---|---|---|---|
| | Responder | 185 | 2.31 | 0.27 | 1.70 | 2.92 |
| | HC | 134 | 2.19 | 0.27 | 1.59 | 2.80 |
| Thalamus | Non-Responder | 52 | 2.09 | 0.23 | 1.10 | 3.07 |
| | Responder | 44 | 1.91 | 0.13 | 1.35 | 2.47 |
| | HC | 32 | 1.95 | 0.14 | 1.37 | 2.54 |
| Basal Ganglia | Non-Responder | 128 | 4.86 | 3.09 | -2.44 | 12.17 |
| | Responder | 101 | 2.37 | 0.89 | 0.27 | 4.46 |
| | HC | 39 | 1.89 | 0.91 | -0.25 | 4.03 |
| **mI** | | | | | | |
| MFC | Non-Responder | 156 | 8.09 | 0.58 | 6.77 | 9.40 |
| | Responder | 171 | 7.59 | 0.58 | 6.28 | 8.90 |
| | HC | 121 | 7.29 | 0.59 | 5.96 | 8.62 |
| Thalamus | Non-Responder | 52 | 4.33 | 0.45 | 2.41 | 6.26 |
| | Responder | 43 | 4.12 | 0.31 | 2.77 | 5.46 |
| | HC | 32 | 4.29 | 0.33 | 2.85 | 5.73 |
| Basal Ganglia | Non-Responder | 130 | 11.06 | 5.98 | -3.09 | 25.20 |
| | Responder | 114 | 6.49 | 1.67 | 2.53 | 10.45 |
| | HC | 26 | 5.69 | 0.27 | 5.04 | 6.33 |
| **GSH** | | | | | | |
| MFC | Non-Responder | 78 | 1.28 | 0.24 | 0.25 | 2.32 |
| | Responder | 55 | 1.31 | 0.24 | 0.28 | 2.35 |
| | HC | 25 | 1.34 | 0.24 | 0.30 | 2.38 |

Abbreviations: Cho: choline; CI: confidence interval; DLPFC: dorsolateral prefrontal cortex; EMMs: estimated marginal means; GABA: gamma-aminobutyric acid; Glu: glutamate; Glx: glutamate + glutamine; GSH: glutathione; HC: healthy controls; mI: myo-inositol; MFC: medial frontal cortex; NAA: N-acetylaspartate; SE: standard error.

### Meta-analysis – Methods

*Data extraction*

Metabolite means and standard deviations of treatment-responders and treatment non-responders with schizophrenia and healthy controls (HC) were extracted independently (B.K, J.S) and then compared. Metabolite data were categorised into the following brain regions: (1) medial frontal cortex (MFC), including voxels in the anterior cingulate cortex (ACC), and mid cingulate cortex (MCC); (2) dorsolateral prefrontal cortex (DLPFC); (3) thalamus; and (4) basal ganglia (including the putamen, caudate and striatum). We also extracted demographic and clinical data including age, sex, antipsychotic medication, medication dose and whether the 'non-responder' group were defined in the manuscript as meeting criteria for TRS (including UTRS). For longitudinal studies, only the metabolite values for the first time point were included. When the same sample or partially overlapping samples were included in more than one report, we included the data from the study with the largest sample or kept both samples if the publications reported on different metabolites. When more than one group of treatment non-responders was reported in a single study, (e.g. TRS and UTRS), each non-responder group was treated as an individual dataset, and the sample size of the comparator groups was divided accordingly. In the primary meta-analyses, if a study reported data from multiple voxels within a single brain region (e.g., pACC and dACC), we included the voxel with the largest sample size to maximise statistical power.

### Meta-analysis - Statistics

*Meta-analyses of mean differences*

As our primary objective was to determine differences between treatment non-responder and responder groups, we conducted pairwise analyses which, in addition to offering simplicity, are advantageous for meta-analysis as they restrict comparisons to direct within-study contrasts (given that not all studies included all three groups). This approach avoids assumptions of transitivity and accounts for the possibility that heterogeneity may differ between group comparisons. Subsequent analysis compared each patient group to healthy volunteers to aid interpretation. Meta-analyses of standardised mean differences (SMDs) between groups were conducted using random-effects models in the "metafor" R package (24). SMDs were calculated using the Hedge's g statistic to adjust for small sample bias. Heterogeneity was measured using the $I^2$ value, with higher percentages indicating higher variation across studies in the meta-analysis. The Benjamini–Hochberg false discovery rate (FDR) procedure was applied to all analyses, with a Q threshold of 10%, excluding comparisons of MFC glutamate, Glx, Cho and mI.

*Meta analyses of variability*

Meta-analyses of variability between groups were quantified using the log coefficient of variation ratios (CVR), as described previously (25,26). The Benjamini–Hochberg false discovery rate (FDR) procedure was applied to all analyses, with a Q threshold of 10%, excluding comparisons involving glutamatergic metabolites in the MFC and basal ganglia.

**Meta-analysis - Included studies**

The initial search identified 1,989 articles, of which 23 met the inclusion criteria (PRISMA flow diagram presented in Supplementary Figure 1; see also Supplementary Table 9).

Across included studies, 16 reported Glu, 16 reported Glx, 14 reported NAA, 12 reported mI, 13 reported Cho, 4 reported GABA, and 3 reported GSH. Spectra were acquired in the medial frontal cortex (MFC; 21 studies), dorsolateral prefrontal cortex (DLPFC; 4 studies), thalamus (5 studies), and basal ganglia (7 studies).

In total, the meta-analysis included 652 individuals classed as treatment responders (mean [range] age, 32 [21–47] years), 601 classed as treatment non-responders (31 [19–47] years), and 591 healthy controls (32 [19–44] years). The mean group sample size (range) was responder group: 21 (5–104), non-responder group: 19 (3–38), and healthy control group 26 (6–117).

**Supplementary Table 9. List of studies included in meta-analyses of mean differences and variability**

| Author (ref) | Year | MRS Voxels | Participant group (N) | Site subgroup | Response group | Correction method | Design | Metabolites |
|---|---|---|---|---|---|---|---|---|
| Egerton (1) | 2012 | MFC<br>Thalamus | FEP (22) | | Remission (15)<br>Non-remission (17) | Cr | Cross-sectional | Glu, Glx, NAA, mI, Cho |
| Szulc (27) | 2013 | Thalamus | Schizophrenia (42)<br>HC (26) | | Responders (17)<br>Non-responders (25) | Cr | Prospective | Glx, NAA, mI, Cho |
| McIlwain (2) | 2015 | MFC<br>DLPFC<br>Basal Ganglia (Putamen) | Schizophrenia (42)<br>HC (16) | | FLO (15)<br>TRS (16)<br>UTRS (11) | Cr | Cross-sectional | Glu, Glx, NAA, Cho |
| Mouchlianitis (3) | 2015 | MFC | Schizophrenia (41) | | Treatment responders (20)<br>TRS (21) | Cr | Cross-sectional | Glu, Glx, NAA, mI, Cho |
| Egerton (4) | 2018 | MFC<br>Thalamus | FEP (71) | Site A | Remission (17)<br>Non-remission (5) | Cr | Prospective | Glu, Glx, NAA, mI, Cho |
| | | | | Site B | Remission (16)<br>Non-remission (13) | | | |
| | | | | Site C | Remission (8)<br>Non-remission (6) | | | |
| Iwata (5) | 2019 | MFC<br>DLPFC<br>Basal Ganglia (caudate) | Schizophrenia (74)<br>HC (26) | | Non-TRS (21)<br>Non-UTRS (27)<br>UTRS (26) | CSF | Cross-sectional | Glu, Glx, NAA, Cho |

| | | | | | | | | |
|---|---|---|---|---|---|---|---|---|
| Bojesen (6) | 2020 | MFC<br>Thalamus | FEP (44)<br>HC (35) | | Responders (11)<br>Non-responders (19) | CSF and Cr | Prospective | Glu, Glx, NAA, mI, Cho, GABA |
| Dempster (7) | 2020 | MFC | FEP (26)<br>HC (27) | | Remission (11)<br>Non-remission (15) | CSF | Prospective | Glu, GSH |
| Li (28) | 2020 | MFC | FEP (35)<br>HC (40) | | Remission (25)<br>Non-remission (10) | CSF and Cr | Prospective | Glu |
| Tarumi (29) | 2019 | MFC<br>Basal Ganglia (caudate) | Schizophrenia (59)<br>HC (29) | | Non-TRS (31)<br>TRS (28) | CSF | Cross-sectional | Glu, Glx, NAA, mI, Cho |
| Egerton (8) | 2021 | MFC<br>Basal Ganglia (striatum) | Schizophrenia (92) | Site A | Responder (16)<br>Non-responder (18) | CSF | Prospective | Glu, Glx, NAA, mI, Cho |
| | | | | Site B | Responder (17)<br>Non-responder (15) | | | |
| | | | | Site C | Responder (8)<br>Non-responder (5) | | | |
| | | | | Site D | Responder (7)<br>Non-responder (6) | | | |
| Horne (9) | 2021 | MFC | Schizophrenia (40)<br>HC (20) | | Responders (21)<br>TRS (19) | Cr | Cross-sectional | Glu, Glx, NAA, mI, Cho |
| Iwata (10) | 2021 | MFC | Schizophrenia (72)<br>HC (26) | | Clz non-responders (24)<br>Clz Responders (27)<br>FLR (21) | CSF | Cross-sectional | GSH |
| Yang (11) | 2021 | MFC<br>DLPFC<br>Thalamus | Schizophrenia (136) | | non-TR (104)<br>TR (32) | CSF and Cr | Cross-sectional | NAA, GABA, GSH |

| | | | | | | | | |
|---|---|---|---|---|---|---|---|---|
| Huang (12) | 2022 | MFC<br>mPFC | Schizophrenia (73)<br>HC (19) | | Non-TRS (35)<br>TRS (38) | NAA ratio | Cross-sectional | Glx |
| Matrone (13) | 2022 | MFC | TRS (20)<br>HC (10) | | TRS (20) | Cr | Cross-sectional | Glu, Glx |
| Reyes-Madrigal (14) | 2022 | Basal Ganglia (striatum) | FEP (48) | | Responders (29)<br>Non-responders (19) | CSF | Prospective | Glu, Glx, NAA, mI, Cho |
| Ueno (15) | 2022 | MFC | Schizophrenia (63)<br>HC (35) | | FLR (16)<br>non-URS (25)<br>URS (22) | CSF and Cr | Cross-sectional | GABA |
| Egerton (16) | 2023 | MFC<br>Basal Ganglia (caudate) | FEP (45) | Site A | Responders (19)<br>Non-responders (10) | CSF and Cr | Prospective | Glu, Glx |
| | | | | Site B | Responders (6)<br>Non-responders (7) | | | |
| Fan (17) | 2023 | MFC (pACC, dACC) | FEP (42)<br>HC (41) | | Responders (27)<br>Non-responders (15) | CSF | Prospective | Glu, Glx, NAA, mI, Cho |
| Van Der Pluijm (18) | 2024 | MFC | FEP (78)<br>HC (20) | | Responders (46)<br>TRS (12) | CSF and Cr | Prospective | Glx, NAA, mI, Cho, GABA |

| | | | | | | | |
|---|---|---|---|---|---|---|---|
| Torres-Carmona (30) | 2024 | MFC<br>DLPFC<br>Basal Ganglia (striatum) | Clz-R (37)<br>Clz-NR (30)<br>Treatment responders (38)<br>HC (52) | Responders (38)<br>Clz-R (TRS) (37)<br>Clz-NR (UTRS) (30) | CSF | Cross-sectional | mI |
| Maximo (31) | 2025 | MFC | FEP (113)<br>HC (117) | Remission (61)<br>Non-remission (23) | CSF | Prospective | Glu |

Note: Where studies acquired data at more than one site, data are treated separately. Abbreviations: Cr: Creatine; Cho: choline; Clz-NR: clozapine non-responders; Clz-R: clozapine responders; DLPFC: dorsolateral prefrontal cortex; FEP: first-episode psychosis; FLR: first line responders; GABA: gamma-aminobutyric acid; Glu: glutamate; Glx: glutamate + glutamine; GSH: glutathione; HC: healthy controls; mI: myo-inositol; MFC: medial frontal cortex; NAA: N-acetylaspartate; TRS: treatment resistant schizophrenia; UTRS: ultra treatment resistant schizophrenia.

**Supplementary Table 10. Summary of results for meta-analysis of mean differences**

| Metabolite | Meta analysis (N participants) | N cohorts | Effect size SMD (Hedge's G) (95% CI) | P value | $I^2$ |
|---|---|---|---|---|---|
| **Glu** | | | | | |
| MFC | NR (349) vs R (464) | 23 | 0.20 (-0.04 to 0.43) | 0.098 | 55 |
| | NR (243) vs HC (416) | 15 | 0.09 (-0.22 to 0.40) | 0.565 | 66 |
| | R (257) vs HC (406) | 14 | 0.07 (-0.14 to 0.28) | 0.501 | 34 |
| DLPFC | NR (102) vs R (138) | 5 | -0.89 (-1.45 to -0.32) | 0.002 | 64 |
| | NR (70) vs HC (42) | 4 | -0.01 (-0.62 to 0.60) | 0.974 | 57 |
| | R (34) vs HC (42) | 4 | 1.18 (0.35 to 2.01) | 0.005 | 63 |
| Thalamus | NR (66) vs R (155) | 5 | 0.16 (-0.14 to 0.46) | 0.307 | 0 |
| | NR (18) vs HC (58) | 3 | 0.17 (-0.36 to 0.70) | 0.532 | 0 |
| | R (38) vs HC (58) | 3 | -0.14 (-0.87 to 0.59) | 0.707 | 62 |
| Basal Ganglia | NR (166) vs R (151) | 12 | 0.01 (-0.22 to 0.24) | 0.937 | 0 |
| | NR (90) vs HC (61) | 5 | 0.20 (-0.13 to 0.54) | 0.227 | 0 |
| | R (59) vs HC (61) | 5 | 0.20 (-0.16 to 0.57) | 0.269 | 0 |
| **Glx** | | | | | |
| MFC | NR (319) vs R (343) | 21 | 0.18 (-0.07 to 0.44) | 0.158 | 56 |
| | NR (245) vs HC (271) | 14 | 0.17 (-0.13 to 0.47) | 0.260 | 58 |
| | R (240) vs HC (261) | 13 | 0.05 (-0.31 to 0.42) | 0.776 | 72 |
| DLPFC | NR (70) vs R (34) | 4 | -1.54 (-2.78 to -0.29) | 0.016 | 85 |
| | NR (70) vs HC (42) | 4 | 0.08 (-0.61 to 0.7) | 0.824 | 67 |
| | R (34) vs HC (42) | 4 | 1.72 (0.41 to 3.02) | 0.010 | 82 |
| Thalamus | NR (59) vs R (70) | 5 | 0.14 (-0.48 to 0.76) | 0.658 | 61 |
| | NR (44) vs HC (83) | 4 | 0.03 (-0.37 to 0.44) | 0.867 | 5 |
| | R (57) vs HC (83) | 4 | 0.01 (-0.42 to 0.44) | 0.968 | 32 |
| Basal Ganglia | NR (165) vs R (151) | 12 | 0.01 (-0.36 to 0.38) | 0.968 | 56 |
| | NR (90) vs HC (61) | 5 | 0.46 (-0.27 to 1.19) | 0.217 | 75 |
| | R (59) vs HC (61) | 5 | 0.32 (-0.05 to 0.68) | 0.088 | 0 |
| **NAA** | | | | | |
| MFC | NR (296) vs R (387) | 19 | 0.15 (-0.02 to 0.32) | 0.077 | 6 |
| | NR (187) vs HC (242) | 12 | 0.15 (-0.23 to 0.54) | 0.435 | 70 |
| | R (205) vs HC (242) | 12 | -0.03 (-0.36 to 0.30) | 0.868 | 61 |
| DLPFC | NR (102) vs R (138) | 5 | 0.23 (-0.50 to 0.96) | 0.538 | 80 |
| | NR (34) vs HC (42) | 4 | -0.09 (-0.86 to 0.69) | 0.826 | 73 |
| | R (70) vs HC (42) | 4 | -0.48 (-1.94 to 0.98) | 0.518 | 88 |
| Thalamus | NR (94) vs R (176) | 6 | -0.09 (-0.37 to 0.20) | 0.558 | 7 |
| | NR (45) vs HC (85) | 4 | -0.24 (-0.62 to 0.14) | 0.212) | 16 |
| | R (58) vs HC (85) | 4 | -0.49 (-1.14 to 0.16) | 0.136 | 57 |

| | | | | | |
|---|---|---|---|---|---|
| Basal Ganglia | NR (150) vs R (132) | 10 | -0.04 (-0.35 to 0.27) | 0.803 | 33 |
| | NR (90) vs HC (61) | 5 | 0.02 (-0.31 to 0.35) | 0.915 | 0 |
| | R (59) vs HC (61) | 5 | 0.06 (-0.30 to 0.42) | 0.750 | 0 |
| **mI** | | | | | |
| MFC | NR (255) vs R (287) | 16 | 0.28 (0.10 to 0.46) | **0.002** | 1 |
| | NR (178) vs HC (256) | 10 | 0.45 (0.07 to 0.83) | **0.022** | 69 |
| | R (209) vs HC (256) | 10 | 0.25 (0.00 to 0.51) | 0.054 | 44 |
| Thalamus | NR (61) vs R (71) | 5 | -0.25 (-0.61 to 0.10) | 0.164 | 0 |
| | NR (44) vs HC (85) | 4 | -0.27 (-0.66 to 0.12) | 0.173 | 0 |
| | R (57) vs HC (85) | 4 | -0.07 (-0.41 to 0.27) | 0.676 | 0 |
| Basal Ganglia | NR (150) vs R (135) | 8 | 0.16 (-0.08 to 0.39) | 0.198 | 0 |
| | NR (90) vs HC (75) | 3 | 0.46 (0.14 to 0.77) | 0.004 | 0 |
| | R (65) vs HC (75) | 3 | 0.31 (-0.03 to 0.64) | 0.074 | 0 |
| **Choline** | | | | | |
| MFC | NR (264) vs R (283) | 18 | 0.35 (0.04 to 0.65) | **0.026** | 63 |
| | NR (187) vs HC (242) | 12 | 0.63 (0.15 to 1.11) | **0.010** | 79 |
| | R (205) vs HC (242) | 12 | 0.45 (0.09 to 0.81) | **0.015** | 67 |
| DLPFC | NR (70) vs R (34) | 4 | 1.05 (0.18 to 1.93) | 0.019 | 73 |
| | NR (70) vs HC (42) | 4 | 1.77 (0.77 to 2.77) | 0.001 | 78 |
| | R (34) vs HC (42) | 4 | 0.68 (0.22 to 1.15) | 0.004 | 0 |
| Thalamus | NR (62) vs R (72) | 5 | -0.13 (-0.48 to 0.23) | 0.481 | 0 |
| | NR (45) vs HC (85) | 4 | -0.36 (-0.75 to 0.03) | 0.070 | 0 |
| | R (58) vs HC (85) | 4 | -0.26 (-0.60 to 0.08) | 0.129 | 0 |
| Basal Ganglia | NR (150) vs R (132) | 10 | 0.02 (-0.30 to 0.35) | 0.885 | 39 |
| | NR (90) vs HC (61) | 5 | 0.04 (-0.60 to 0.60) | 0.907 | 69 |
| | R (59) vs HC (61) | 5 | -0.03 (-0.52 to 0.47) | 0.919 | 39 |
| **GABA** | | | | | |
| MFC | NR (102) vs R (173) | 5 | 0.02 (-0.26 to 0.31) | 0.886 | 3 |
| | NR (70) vs HC (85) | 4 | -0.26 (-0.70 to 0.17) | 0.237 | 41 |
| | R (69) vs HC (85) | 4 | -0.19 (-0.54 to 0.17) | 0.303 | 0 |
| **GSH** | | | | | |
| MFC | NR (94) vs R (135) | 4 | -0.25 (-0.68 to 0.18) | 0.258 | 45 |
| | NR (62) vs HC (51) | 3 | 0.06 (-0.33 to 0.45) | 0.753 | 0 |
| | R (31) vs HC (51) | 3 | 0.11 (-0.34 to 0.57) | 0.620 | 0 |

Note: Standardised mean differences (SMDs), Hedges' g with 95% confidence intervals (95% CI). Significant results are indicated in bold. Abbreviations: Cho: choline; DLPFC: dorsolateral prefrontal cortex; GABA: gamma-aminobutyric acid; Glu: glutamate; Glx: glutamate+glutamine; GSH: glutathione; HC: healthy controls; MFC: medial frontal cortex; mI: myo-inositol; NAA: N-acetylaspartate; NR: non-responder group; R: responder group.

### Supplementary Figure 3

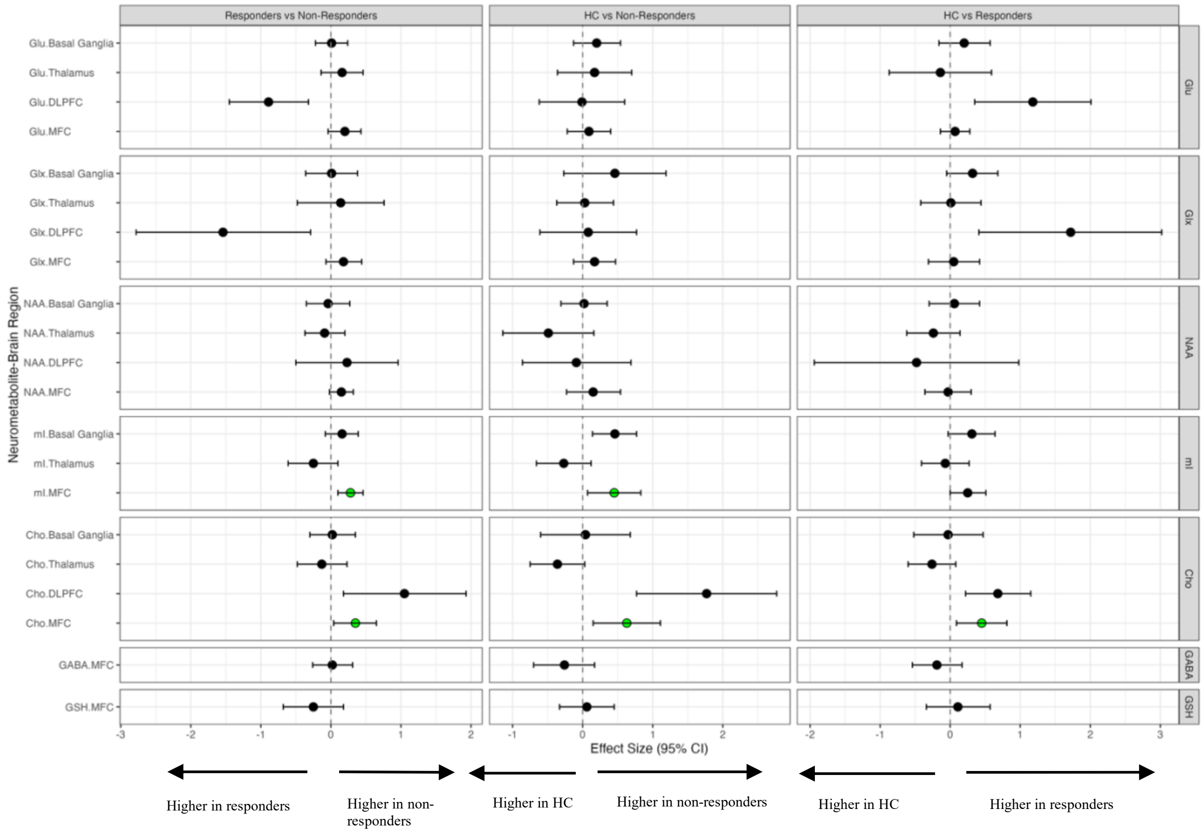


**Forest plot showing Hedge's G effect sizes for standardised mean differences of [1]H-MRS metabolites.**

Significant results (after FDR correction as described in the methods) are shown in green. Abbreviations: Cho: choline; DLPFC: dorsolateral prefrontal cortex; GABA: gamma-aminobutyric acid; Glu: glutamate; Glx: glutamate+glutamine; GSH: glutathione; MFC: medial frontal cortex; mI: myo-inositol; NAA: N-acetylaspartate.

**Supplementary Table 11. Meta analysis of variability (CVR)**

| Metabolite | Meta analysis (N participants) | N cohorts | Effect size SMD (95% CI) | P value | I2 |
|---|---|---|---|---|---|
| **Glu** | | | | | |
| MFC | NR (349) vs R (464) | 23 | 0.01 (-0.11 to 0.13) | 0.877 | 37 |
| | NR (243) vs HC (416) | 17 | 0.15 (0 to 0.31) | 0.046 | 55 |
| | R (257) vs HC (406) | 16 | 0.08 (-0.04 to 0.21) | 0.197 | 40 |
| DLPFC | NR (102) vs R (138) | 5 | -0.21 (-0.38 to -0.03) | 0.024 | 0 |
| | NR (70) vs HC (42) | 4 | 0.16 (-0.09 to 0.41) | 0.202 | 0 |
| | R (34) vs HC (42) | 4 | -0.08 (-0.39 to 0.22) | 0.598 | 0 |
| Thalamus | NR (66) vs R (155) | 5 | 0.06 (-0.29 to 0.42) | 0.724 | 58 |
| | NR (18) vs HC (58) | 4 | -0.16 (-0.85 to 0.53) | 0.645 | 73 |
| | R (38) vs HC (58) | 4 | 0.07 (-0.2 to 0.34) | 0.611 | 0 |
| Basal Ganglia | NR (166) vs R (151) | 12 | -0.06 (-0.28 to 0.16) | 0.606 | 54 |
| | NR (90) vs HC (61) | 5 | -0.2 (-0.45 to 0.05) | 0.115 | 43 |
| | R (59) vs HC (61) | 5 | -0.23 (-0.43 to -0.02) | 0.032 | 0 |
| **Glx** | | | | | |
| MFC | NR (319) vs R (343) | 21 | -0.12 (-0.28 to 0.05) | 0.165 | 64 |
| | NR (245) vs HC (271) | 17 | 0.24 (0.11 to 0.38) | 0.000 | 40 |
| | R (240) vs HC (261) | 16 | -0.02 (-0.2 to 0.16) | 0.791 | 71 |
| DLPFC | NR (70) vs R (34) | 4 | -0.24 (-0.51 to 0.03) | 0.084 | 0 |
| | NR (70) vs HC (42) | 4 | 0.21 (-0.03 to 0.46) | 0.092 | 0 |
| | R (34) vs HC (42) | 4 | -0.03 (-0.33 to 0.27) | 0.844 | 0 |
| Thalamus | NR (59) vs R (70) | 5 | -0.19 (-0.48 to 0.09) | 0.189 | 0 |
| | NR (44) vs HC (83) | 5 | 0.31 (0.04 to 0.58) | 0.024 | 0 |
| | R (57) vs HC (83) | 5 | 0.03 (-0.22 to 0.29) | 0.797 | 4 |
| Basal Ganglia | NR (165) vs R (151) | 12 | 0 (-0.15 to 0.16) | 0.952 | 0 |
| | NR (90) vs HC (61) | 5 | -0.11 (-0.36 to 0.14) | 0.387 | 22 |
| | R (59) vs HC (61) | 5 | -0.05 (-0.25 to 0.16) | 0.668 | 0 |
| **NAA** | | | | | |
| MFC | NR (296) vs R (387) | 19 | 0.03 (-0.13 to 0.19) | 0.699 | 52 |
| | NR (187) vs HC (242) | 14 | 0.27 (0.15 to 0.39) | 0.000 | 0 |
| | R (205) vs HC (242) | 14 | 0.1 (-0.07 to 0.27) | 0.260 | 56 |
| DLPFC | NR (102) vs R (138) | 5 | -0.07 (-0.26 to 0.12) | 0.459 | 0 |
| | NR (70) vs HC (42) | 4 | 0.14 (-0.11 to 0.39) | 0.278 | 0 |
| | R (34) vs HC (42) | 4 | 0.02 (-0.29 to 0.33) | 0.899 | 0 |
| Thalamus | NR (94) vs R (176) | 6 | -0.14 (-0.37 to 0.1) | 0.255 | 36 |
| | NR (45) vs HC (85) | 5 | 0.18 (-0.11 to 0.47) | 0.228 | 29 |

| | | | | | |
|---|---|---|---|---|---|
| | R (58) vs HC (85) | 5 | -0.01 (-0.21 to 0.19) | 0.943 | 0 |
| Basal Ganglia | NR (150) vs R (132) | 10 | 0.13 (-0.24 to 0.5) | 0.489 | 75 |
| | NR (90) vs HC (61) | 5 | 0.43 (0.07 to 0.8) | 0.020 | 63 |
| | R (59) vs HC (61) | 5 | 0.48 (0.24 to 0.71) | 0.000 | 0 |
| **mI** | | | | | |
| MFC | NR (255) vs R (287) | 16 | 0.11 (-0.05 to 0.27) | 0.195 | 53 |
| | NR (178) vs HC (256) | 12 | 0.05 (-0.1 to 0.21) | 0.519 | 51 |
| | R (209) vs HC (256) | 12 | 0.06 (-0.04 to 0.17) | 0.229 | 5 |
| Thalamus | NR (61) vs R (71) | 5 | 0.13 (-0.13 to 0.39) | 0.317 | 0 |
| | NR (44) vs HC (85) | 5 | -0.12 (-0.43 to 0.19) | 0.440 | 33 |
| | R (57) vs HC (85) | 5 | -0.07 (-0.32 to 0.17) | 0.544 | 0 |
| Basal Ganglia | NR (150) vs R (135) | 8 | -0.01 (-0.3 to 0.27) | 0.918 | 58 |
| | NR (90) vs HC (75) | 3 | 0.14 (-0.04 to 0.33) | 0.133 | 0 |
| | R (65) vs HC (75) | 3 | 0.12 (-0.09 to 0.33) | 0.272 | 0 |
| **Choline** | | | | | |
| MFC | NR (246) vs R (267) | 17 | 0.04 (-0.11 to 0.2) | 0.587 | 40 |
| | NR (187) vs HC (242) | 14 | 0.01 (-0.17 to 0.18) | 0.922 | 55 |
| | R (205) vs HC (242) | 14 | 0.16 (-0.15 to 0.46) | 0.320 | 78 |
| DLPFC | NR (70) vs R (34) | 4 | 0.11 (-0.16 to 0.37) | 0.419 | 0 |
| | NR (70) vs HC (42) | 4 | -0.04 (-0.29 to 0.2) | 0.725 | 0 |
| | R (34) vs HC (42) | 4 | 0.06 (-0.23 to 0.36) | 0.671 | 0 |
| Thalamus | NR (62) vs R (72) | 5 | 0.1 (-0.28 to 0.48) | 0.618 | 63 |
| | NR (45) vs HC (85) | 5 | -0.01 (-0.29 to 0.27) | 0.924 | 37 |
| | R (58) vs HC (85) | 5 | -0.22 (-0.45 to 0.02) | 0.071 | 23 |
| Basal Ganglia | NR (150) vs R (132) | 10 | 0.24 (-0.06 to 0.54) | 0.116 | 65 |
| | NR (90) vs HC (61) | 5 | -0.04 (-0.33 to 0.25) | 0.800 | 40 |
| | R (59) vs HC (61) | 5 | 0.02 (-0.21 to 0.24) | 0.893 | 0 |
| **GABA** | | | | | |
| MFC | NR (102) vs R (173) | 5 | 0.15 (-0.17 to 0.47) | 0.364 | 62 |
| | NR (70) vs HC (85) | 7 | 0.02 (-0.14 to 0.17) | 0.822 | 7 |
| | R (69) vs HC (85) | 7 | 0.23 (-0.06 to 0.52) | 0.126 | 66 |
| **GSH** | | | | | |
| MFC | NR (94) vs R (135) | 4 | 0.02 (-0.27 to 0.31) | 0.900 | 63 |
| | NR (62) vs HC (51) | 3 | -0.09 (-0.51 to 0.34) | 0.691 | 76 |
| | R (31) vs HC (51) | 3 | -0.13 (-0.43 to 0.18) | 0.422 | 0 |

Note: Coefficient of variation ratio (CVR), 95% confidence intervals (95% CI) P value and $I^2$ are presented. Significant results are indicated in bold. Abbreviations: Cho: choline; DLPFC: dorsolateral prefrontal cortex; Glu: glutamate; Glx: glutamate+glutamine; GSH: glutathione; HC: healthy controls; MFC: medial frontal cortex; mI: myo-inositol; NAA: N-acetylaspartate; NR: non-responder group; R: responder group.

### Supplementary Figure 4

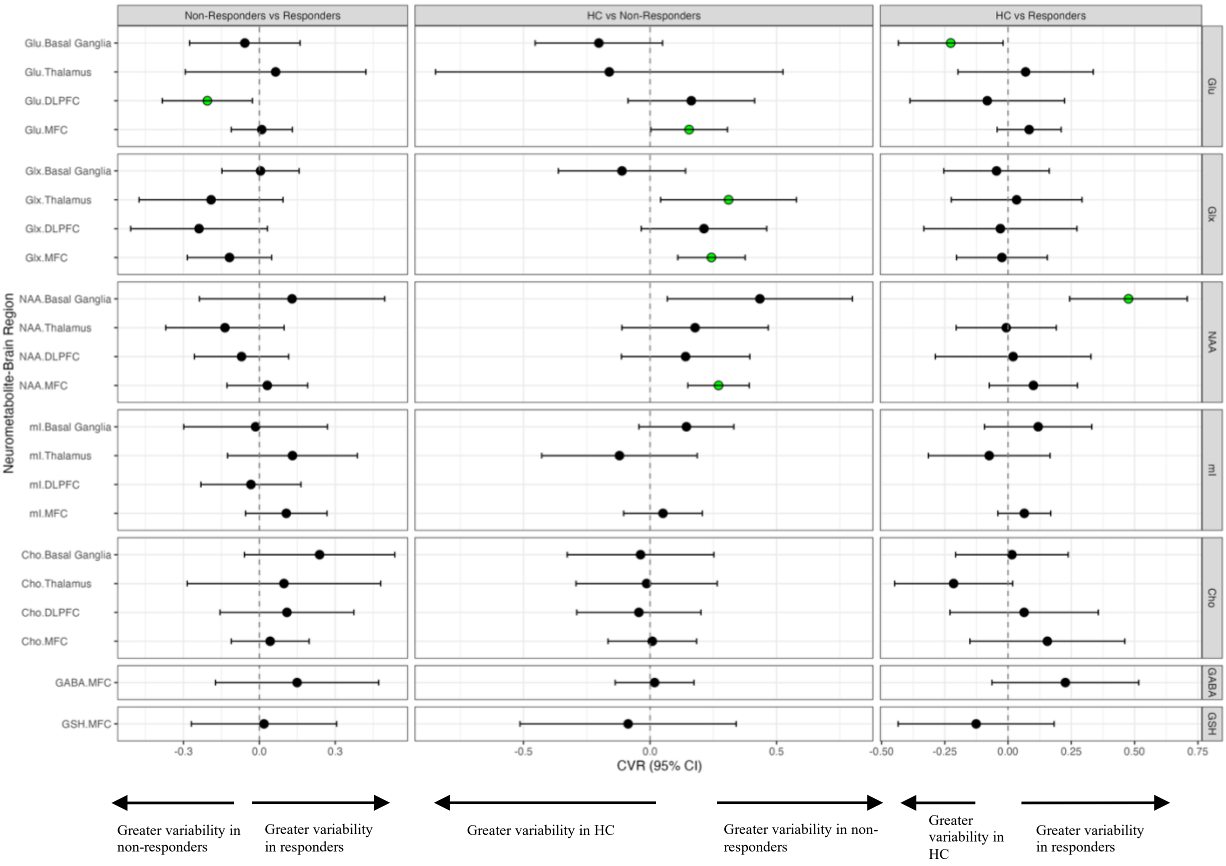


**Forest plot showing the summary effect sizes for the coefficient of variation ratio (CVR) of $^{1}$H-MRS metabolites.**

Significant results (after FDR correction as described in the methods) are shown in green. Abbreviations: Cho: choline; DLPFC: dorsolateral prefrontal cortex; Glu: glutamate; Glx: glutamate+glutamine; GSH: glutathione; MFC: medial frontal cortex; mI: myo-inositol; NAA: N-acetylaspartate.
